# Investigating risks for human colonisation with extended spectrum beta-lactamase producing *E. coli* and *K. pneumoniae* in Malawian households: a one health longitudinal cohort study

**DOI:** 10.1101/2022.08.16.22278508

**Authors:** Derek Cocker, Kondwani Chidziwisano, Madalitso Mphasa, Taonga Mwapasa, Joseph M. Lewis, Barry Rowlingson, Melodie Sammarro, Winnie Bakali, Chifundo Salifu, Allan Zuza, Mary Charles, Tamandani Mandula, Victor Maiden, Stevie Amos, Shevin T Jacob, Henry Kajumbula, Lawrence Mugisha, David Musoke, Rachael Byrne, Thomas Edwards, Rebecca Lester, Nicola Elviss, Adam Roberts, Andrew C Singer, Christopher Jewell, Tracy Morse, Nicholas A Feasey

## Abstract

**Background:** Low- and middle-income countries (LMICs) have high morbidity and mortality from drug-resistant infections, especially from enteric bacteria such as *Escherichia coli*. LMICs have varying infrastructure and services in the community to separate people from human and animal waste, creating risks for ESBL-Enterobacterales (ESBL-E) transmission. Limited data exist from Southern Africa on the prevalence of ESBL-E the community.

**Methods and findings:** In this longitudinal cohort study we took a one-health approach to investigating prevalence and distribution of ESBL-E in urban, peri-urban and rural Malawian households between May 2018 and October 2020. We described human health, antibiotic usage (ABU), health seeking behaviour, structural and behavioural environmental health practices, and animal husbandry at these households. In parallel, human and animal stool and diverse environmental samples were collected and cultured to identify presence of ESBL *E. coli* and ESBL *K. pneumoniae*. Univariable and multivariable analysis was performed to determine associations with human ESBL-E colonisation.

We recruited 300 households, totalling 841 visits, and a paucity of environmental health infrastructure and materials for safe sanitation was noted across all sites. In total, 11,975 samples were cultured and ESBL-E were isolated from 41.8% (n=1190) of human stool and 29.8% (n=290) of animal stool samples. Animal species with particularly high rates of ESBL-E colonisation included pigs (56.8%, n=21) poultry (32.5%, n=148) and dogs (58.8% n= 30). ESBL-E were isolated from 66.2% (n=339) of river water samples and 46.0% (n=138) of drain samples. Urban areas had greater ESBL-E contamination of food, household surfaces, floors and the external environment, alongside the highest rates of ESBL-E colonisation in humans (47.1%, n=384) and animals (55.1%, n=65). Multivariable models illustrated that human ESBL *E. coli* colonisation was associated with the wet season (aOR = 1.66, 95%CrI: 1.38-2.00), living in urban areas (aOR = 2.01, 95%CrI: 1.26-3.24), advanced age (aOR = 1.14, 95%CrI: 1.05-1.24) and in households where animals were observed interacting with food (aOR = 1.62, 95%CrI: 1.17-2.28) or kept inside (aOR = 1.58, 95%CrI: 1.00-2.43). Human ESBL *K. pneumoniae* colonisation was also associated with the wet season (aOR = 2.23, 95%CrI: 1.63-2.76.

**Conclusion:** We identified extremely high levels of ESBL-E colonisation in humans and animals and contamination of the environment in Southern Malawi. Urbanisation and season are key risks for ESBL-E colonisation, perhaps reflecting environmental contamination as toilets overflow in high population density areas in heavy rains in the wet season. Without adequate efforts to improve environmental health, ESBL transmission is likely to persist in this setting.

## Introduction

In sub-Saharan Africa (sSA) there is a high morbidity and mortality from infections caused by antimicrobial resistant (AMR) bacteria, especially extended-spectrum ß-lactamase (ESBL) producing Enterobacterales (1). Given the heavy reliance on 3^rd^ -generation cephalosporins (3GCs) in human health, two of the most important AMR bacteria found in sSA include *Escherichia coli*, responsible for a spectrum of community acquired infections and *Klebsiella pneumoniae*, more typically associated with healthcare associated infection (HCAI) (2). These bacteria are present in the guts of humans and animals and also within the broader environment (3). Households are therefore a focal point from which these enteric bacteria can disseminate via human and animal waste into the environment, potentially facilitating onward transmission of these bacteria to further human and animal hosts (4,5).

In low- and middle-income countries (LMICs), paucity of infrastructure and services to support environmental health (including water, sanitation, food safety and hygiene) are a key facilitator of unrestricted interaction between people and both human and animal waste in the environment. These infrastructural and service delivery inadequacies are compounded by poor hygiene practices, which increase the complexity and opportunity for these interactions (6,7). Environmental health factors are therefore thought to play a central role in environmental ESBL-Enterobacterales (ESBL-E) transmission, which may lead to onward risks for vulnerable individuals (6). Interventions to interrupt community ESBL-E transmission need to target key transmission routes, yet context specific data to guide such interventions are lacking.

It is likely that transmission routes are heterogeneous across different settings: environmental health infrastructure and practices typically differ between urban and rural settings, with urban areas considered at particular risk of AMR transmission due to high-density housing, increased antibiotic use (ABU) and a paucity of environmental health infrastructure (8). Regional differences in animal ownership and husbandry practices are likely to further impact risks of AMR transmission. Therefore, a one health approach interrogating human, animal and environmental health factors across urban and rural settings in LMIC is critical to generate data to inform cost-effective interventions. To date, little evidence exists in the literature on the prevalence of ESBL bacteria in LMIC households and communities, especially one health data incorporating contemporaneously collected data on the prevalence of ESBL colonisation in co-located animals and the local environment (9).

Here, we have placed households in urban, peri-urban and rural settings in Malawi at the heart of our study; our objectives were i) to describe the prevalence of ESBL-E found in human, animal and environmental compartments in Blantyre, Malawi, and ii) identify key one health factors associated with human ESBL-E colonisation to inform future interventions.

## Methods

Between May 2018 and October 2020 we aimed to recruit 300 households, 100 in each of Ndirande (urban), Chileka (peri-urban) and Chikwawa (rural) in the Southern region of Malawi using GPS coordinates derived via an inhibitory with close pair spatial design to avoid systematic biases (see (10) for detailed protocol). These study sites were included to enable variations in environmental health practices, animal practices, antibiotic usage (ABU), and contamination with ESBL-producing bacteria to be contrasted. Households identified at or near GPS locations were screened for inclusion and excluded if (i) they did not fall into the demarcated study boundaries, (ii) had <2 people inhabiting the household, (iii) did not speak English or Chichewa, or (iv) if they did not consent to take part in the study. 65 households per region were assigned for longitudinal follow-up and had 4 visits in total over a 6-month period and the remaining 35 households had a baseline visit alone.

Case report forms (CRFs) were completed at each visit, providing information at both an individual and household level on human health, antibiotic usage (ABU), health seeking behaviour, structural and behavioural environmental health proxies and animal husbandry. In parallel, observational checklists were completed, documenting key environmental health and household sanitation practices. Lastly, at each visit, up to 20 microbiological samples were taken, inclusive of human stool, animal stool and a broad range of environmental sites (See (10) for details).

Samples were incubated in an enrichment broth (buffered peptone water) at 37 ± 1 °C for 18-24 hours and plated onto CHROMagar™ ESBL chromogenic agar (CHROMagar™, France). Plates were then placed in an aerobic incubator at 37 ± 1 °C for 18-24 hours and read for growth of ESBL bacteria. Pink colonies and indole positive white colonies were categorised as ESBL *E. coli*, while blue colonies underwent speciation for *K. pneumoniae* using high resolution melt-curve (HRM) PCR, to identify ESBL *K. pneumoniae* isolates (11).

Statistical analyses and graphic visualisations were performed using R v4.1.2 (R foundation for statistical computing, Vienna, Austria). Summaries are presented as proportions medians +/- interquartile range (IQR) or means +/- standard deviation (SD). Kruskal-Wallis and Fisher’s exact were used to test the equivalence of regional groups (i.e. urban, peri-urban and rural) for continuous and categorical variables respectively. Chi-squared was used to test for differences in bacterial species composition of samples and seasonal variations in prevalence (wet/dry). Wet season was classified as samples obtained between Nov-Apr and dry season was classified as samples obtained between May-Oct.

Statistical analysis used Bayesian logistic regression to identify factors associated with human ESBL-E colonisation; nonindependence of within-participant and within-household samples was accounted for using hierarchical random effects. Before building regression models, principal component analysis (PCA) was used to visualise variation in the dataset across regions (urban, peri-urban, and rural) using the FactoMineR package in R (12). Putative individual-level variables (e.g age, sex), household level variables (e.g. household size, presence of toilet) and environmental contamination variables (e.g. presence of ESBL-E in drain or stored water) likely to be associated with human ESBL-E colonisation were identified *a priori* by the DRUM consortium (**S1a-1c Tables**) and PCA performed on each group of variables, after log-transforming continuous variables. Individuals and households were then plotted in PCA space for each of the groups of variables with 95% confidence ellipses for each region (i.e. the region that contains 95% of all samples that can be drawn from the underlying normal distribution).

A variable selection strategy was used to construct the logistic regression models. The outcome variable was ESBL-E colonisation in human stool, with separate models fit for ESBL *E. coli* and ESBL *K. pneumoniae*. Individual and household variables were considered for inclusion. Environmental contamination variables were not included as these were considered to be on the causal pathway from other variables to the outcome. A stratified univariable analysis using logistic regression in each region separately was performed to determine which variables to include in the final analysis. Variables that were significantly associated with ESBL colonisation by univariable analysis (p<0.05) in any region were considered for inclusion into multivariable models, and those which were not significant or where data was unavailable for at least one region were not included. Region as well as a random intercept per individual, which was nested within a random intercept per household were included in the final models as well as the other selected variables. The models were fit with Stan v2.21.0 via the R *brms* v2.13.5 package with 4 chains per dataset each with 2000 iterations in total, with 1000 warm up iterations. Default priors were used and convergence of models assessed by inspection of traceplots and by the R-hat convergence diagnostic value being close to 1. Outputs were expressed as odds ratios (OR) with 95% credible interval (CrI).

To allow for the possibility that some variables may have different effects on human ESBL-E colonisation in different settings, (for example be protective in urban but not rural settings) further models were built allowing for interaction effects between region and covariates. Here, ESBL *E. coli* and ESBL *K. pneumoniae* univariable models with and without a region by covariate interaction term were fitted and compared by likelihood ratio testing. If the model with the interaction term was a better fit to the data (defined as p < 0.05) then the interaction term was included in the final “regional” model; otherwise it was omitted. These models included the variables selected, alongside a random intercept per individual, nested within a random intercept per household as before, and fit using the same methods outlined in the final model. Households that did not undergo follow-up only provided baseline data, and longitudinal sample data for these households was not included in the final models. A STROBE statement checklist **(S2 Table)** and primary datasets (**S3a-d Tables**) have been included in the supplementary material. Anonymised IDs have been used for participant and households.

Ethical approval was obtained from Liverpool School of Tropical Medicine (LSTM) Research and Ethics Committee, UK (REC, #18-090) and College of Medicine Research and Ethics Committee, Malawi (#P.11/18/2541). In addition, administrative permissions were granted from community leaders and support obtained from local community advisory groups. Sensitizations of study communities were conducted prior to initiation, and full informed written consent was obtained from all household participants recruited into the study, in their local language.

## Results

Between May 2018 and October 2020, 611 households (urban=263, peri-urban=229 and rural=119) were screened and 300 households (100 per region) were recruited (**Fig 1**). 179 households underwent longitudinal visits, 105 underwent a baseline visit only and 16 households were lost to follow-up (841 visits). There were 1351 household members across 300 households, 71.4% (n=965/1351) of whom consented to recruitment.

**Figure 1.**
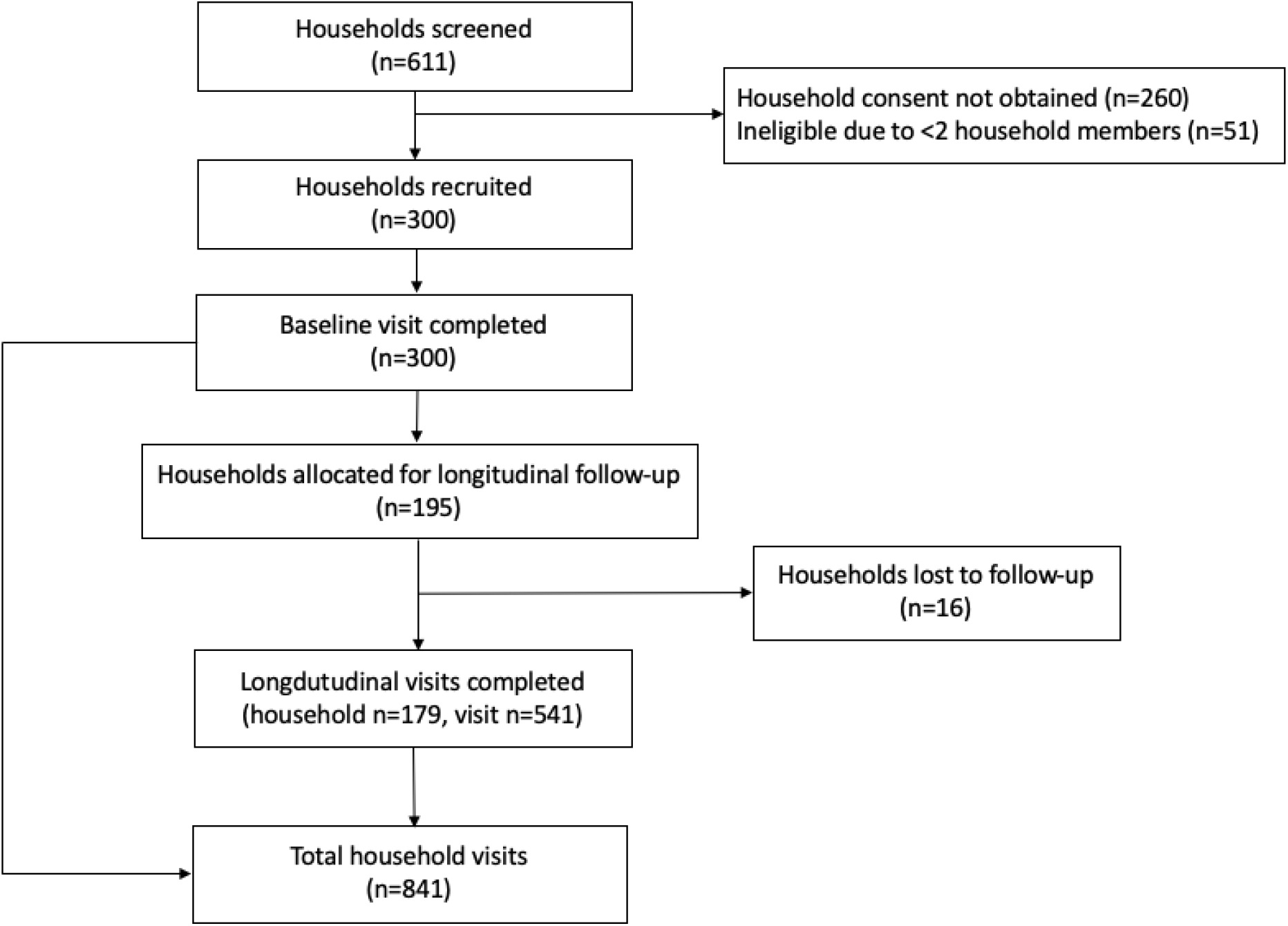
CONSORT chart for households recruited into the study, describing the structure of household visits and loss to follow-up.

### Demographic and human health

The median (IQR) residents per household was 4 (3-5), with urban, peri-urban and rural sites having 4 (3-5), 4 (3-6) and 4 (3-5) members respectively (**Table 1**). Most households comprised of a mix of adults and children, with 2 (2-3) adults, 1 (0-2) adolescent, 0 (0-1) children and 0 (0-1) infants per household. Household income was higher in urban and peri-urban regions than the rural region. However, 97.7% (n=293) of households lived in absolute poverty, as defined by the World Bank (<$1.90/day per individual).

**Table 1.** Baseline household and participant characteristics, stratified by region

| *Household characteristic* | median (IQR) |  |  |  | *p* |
| --- | --- | --- | --- | --- | --- |
|  | Total<br>(*n*=300) | Urban<br>(*n*=100) | Peri-urban<br>(*n*=100) | Rural<br>(*n*=100) |  |
| Number of household members | 4 (3-5) | 4 (3-5) | 4 (3-6) | 4 (3-5) | .281 |
| Household Income (Malawi Kwacha/month)* | 30,000<br>(20,000-50,000) | 50,000<br>(28,750-60,000) | 40,000<br>(30,000-70,000) | 20,000<br>(10,000-30,000) | >.001 |
| *Individual characteristics* | *n (%) unless otherwise stated* |  |  |  | *p* |
|  | Total<br>(*n*=965) | Urban<br>(*n*=312) | Peri-urban<br>(*n*=383) | Rural<br>(*n*=270) |  |
| Age [median yrs (IQR)] | 18 (7-34) | 15 (7-32) | 20 (9-37) | 17 (7-32) | .031 |
| Sex, male | 420 (43.5%) | 122 (39.1%) | 170 (44.4%) | 128 (47.4%) | .121 |
| Job status |  |  |  |  |  |
| Student | 372 (38.5%) | 137 (43.9%) | 140 (36.6%) | 95 (35.2%) | .059 |
| Unemployed | 379 (39.3%) | 107 (34.3%) | 129 (33.7%) | 143 (53.0%) | >.001 |
| Employed | 214 (22.2%) | 68 (21.8%) | 114 (29.7%) | 32 (11.8%) |  |
| *Health status and healthcare exposures* |  |  |  |  |  |
| Co-morbidities, yes | 66 (6.8%) | 12 (4.0%) | 24 (6.3%) | 30 (11.1%) | .003 |
| Living with HIV | 66 (14.0%) | 19 (10.5%) | 26 (23.6%) | 21 (11.3%) | .026 |
| Previous TB | 12 (1.2%) | 5 (2.0%) | 4 (1.0%) | 3 (1.1%) | .765 |
| Illness episode in last month | 154 (16.0%) | 35 (11.2%) | 65 (17.0%) | 54 (20.0%) | .011 |
| Healthcare exposure as patient (last 6 months) | 25 (2.6%) | 6 (1.9%) | 10 (2.6%) | 9 (3.3%) | .541 |
| Healthcare exposure as guardian (last 6 months) | 28 (2.9%) | 6 (1.9%) | 8 (2.1%) | 14 (5.2%) | .046 |
| Healthcare exposure for work | 4 (0.4%) | 2 (0.6%) | 1 (0.3%) | 1 (0.4%) | .828 |
| *Medication usage<sup>^</sup>* |  |  |  |  |  |
| Non-communicable disease medications | 28 (2.9%) | 12 (3.8%) | 6 (1.6%) | 10 (3.7%) | .120 |
| Antiretroviral therapy* | 62 (93.9%) | 17 (89.5%) | 25 (96.2%) | 20 (95.2%) | .263 |
| Co-trimoxazole preventative therapy* | 60 (90.9%) | 16 (84.2%) | 24 (92.3%) | 20 (95.2%) | .105 |
| *Antibiotic exposure in last 6 months* |  |  |  |  |  |
| Antibiotic usage, yes | 147 (15.2%) | 51 (16.3%) | 35 (9.1%) | 61 (22.6%) | >.001 |

\* Adjusted for HIV individuals in each site (urban (*n*=19), peri-urban (*n*=26) and rural (*n*=21)). <sup>^</sup>Ongoing at baseline. \*1000MK = ~1 US Dollar.

The median age of the study population was 18 years (IQR 7-34), and 56.5% (n=545) of household respondents were women (**Table 1**). 51% (n=492) of the study population had no documented HIV status, and the HIV prevalence amongst those with a reported HIV test was 14.0% (n=66), highest in the peri-urban region, with high anti-retroviral therapy (ART) and co-trimoxazole preventative therapy (CPT) uptake (93.9% [n=62] on ART and 90.9% [n=60] on CPT).

There were no cases of active TB in the cohort, and only 12 participants reported past TB therapy. Non-infectious comorbidities were infrequent, with 6.8% (n=66) of the cohort reporting conditions such as hypertension, peptic ulcer disease and COPD, and only 2.9% (n=28) took any form of regular medication other than ART or CPT (**Table 1**). There were low levels of recent healthcare exposure, with 2.6% (n=25) of participants admitted to hospital, 2.9% of participants attending hospital as a guardian and 0.4% of participants working in healthcare settings during the last 6 months, although 15.2% (n=142) of participants received antibiotics in the last 6 months (**Table 1**) predominantly limited to oral amoxicillin, metronidazole and co-trimoxazole (**S4a Table**). Children were more likely to be prescribed antibiotics, with under 5s being the age group with the greatest chance of having been given an antibiotic in the last 6 months (**S4b Table**).

### Animal husbandry practices

58.7% (n=176) of households reported co-habitation with domestic or livestock animals, with 36.0% (n=36), 59.0% (n=59) and, 81.0% (n=81) of households in the urban, peri-urban and rural sites owning ≥1 animal respectively **(Table 2)**. A total of n=2169 animals were linked to a study household at baseline, and both the composition of species and number of animals present per households varied by region **(Table 2 and S5 Table)**. Companion animals (cats and dogs) were located in low numbers per house and made up a large proportion of the animal species owned in urban (n=23/36, 63.9%) and peri-urban (n=25/59, 42.4%) households. Many households kept poultry (i.e. chickens, doves and ducks), and chickens were both the most commonly owned and most numerous animals (urban=18% (n=18), peri-urban=39% (n=39) and rural=59% (n=59) households). Larger animals (pigs, goats and cattle) were seen at fewer households and primarily in the rural or peri-urban settings, highlighting regional differences in animal ownership. 25.7% of households specifically owned animals for breeding and selling purposes, especially in the rural area.

**Table 2.** Baseline animal husbandry characteristics, stratified by region

|  | n (%) |  |  |  | p |
| --- | --- | --- | --- | --- | --- |
|  | Total<br>(n=300) | Urban<br>(n=100) | Peri-urban<br>(n=100) | Rural<br>(n=100) |  |
| **Animal husbandry** |  |  |  |  |  |
| Households that own animals | 176 (58.7%) | 36 (36%) | 59 (59%) | 81 (81%) | > .001 |
| Species of animals owned<sup>^</sup> |  |  |  |  |  |
| Chickens | 116 (38.7%) | 18 (18.0%) | 39 (39.0%) | 59 (59.0%) | > .001 |
| Goats | 49 (16.3%) | 0 (0.0%) | 12 (12.0%) | 37 (37.0%) | > .001 |
| Dogs | 43 (14.3%) | 14 (14.0%) | 19 (19.0%) | 10 (10.0%) | .212 |
| Cats | 24 (8.0%) | 9 (9.0%) | 6 (6.0%) | 7 (7.0%) | .790 |
| Cattle | 23 (7.7%) | 0 (0.0%) | 0 (0.0%) | 23 (23.0%) | NA |
| Pigs | 17 (5.7%) | 0 (0.0%) | 5 (5.0%) | 12 (12.0%) | > .001 |
| Ducks | 14 (4.7%) | 2 (2.0%) | 2 (2.0%) | 10 (10.0%) | .017 |
| Doves | 13 (4.3%) | 1 (1.0%) | 5 (5.0%) | 7 (7.0%) | .092 |
| Other | 6 (2.8%) | 2 (2.4%) | 3 (3.5%) | 1 (2.4%) | 1.00 |
| Guinea fowl | 3 (1.0%) | 0 (0.0%) | 0 (0.0%) | 3 (3.0%) | .109 |
| Turkeys | 2 (0.7%) | 0 (0.0%) | 2 (2.0%) | 0 (0.0%) | .331 |
| Households that rear animals to sell? | 77 (25.7%) | 3 (3.0%) | 25 (25.0%) | 49 (49.0%) | > .001 |
| Chickens kept inside the house, yes* | 70 (60.3%) | 15 (83.3%) | 25 (64.1%) | 30 (50.8%) | .041 |
| Goats kept inside the house, yes* | 8 (16.3%) | NA | 5 (41.7%) | 3 (8.1%) | .015 |
| **Animal disease and antibiotic usage** |  |  |  |  |  |
| Disease noted in last 12 months, yes |  |  |  |  |  |
| Poultry | 51 (44.3%) | 8 (44.4%) | 17 (44.7%) | 26 (44.0%) |  |
| Cattle | 8 (34.8%) | NA | NA | 8 (34.8%) |  |
| Pigs | 4 (23.5%) | NA | 0 (0.0%) | 4 (33.3%) |  |
| Goats | 11 (22.4%) | NA | 1 (8.3%) | 10 (27.0%) |  |
| Access to professional animal health services | 47 (26.9%) | 7 (19.4%) | 11 (19.0%) | 29 (35.8%) | 0.054 |
| Antibiotics given to animals in the last 2 months | 7 (4.0%) | 1 (2.8%) | 2 (3.4%) | 4 (4.9%) | 1.00 |

\*= denominator includes only those households that either owned chickens or goats.

Regional differences in animal husbandry practices were found, with animals frequently kept inside the house in the urban setting, particularly poultry (**Table 2 & S5 Table)**. Co-located animals often had episodes of presumed illness, especially poultry (43.3%, n=51), but households had limited access to, or awareness of, veterinarian services (26.9%, n=47). Households reported that they would often do nothing if animals became unwell (**S6 Table**), and only 7 households treated any animals with antibiotics prior to recruitment into the study (**Table 2**).

### Environmental interaction and environmental health

Households typically obtained water from boreholes (48.7%, n=153), public kiosks (25.2%, n=79) or taps piped into the household compound (16.9%, n=53). Water was infrequently treated prior to drinking (8.3%, n=25) and often left uncovered when stored (31.9%, n=143). In total, 89.0% (n=267) of households owned a toilet, most commonly a pit latrine (88.8%, n=137), and frequently shared their toilet with other non-household members (41.9%, n=112). Open defecation was common, with 28.7% (n=86) of households reporting at least 1 member of the household practicing open defecation, and human faeces were found on the floor in or around the household compound at 8.1% of visits. Access to infrastructure and consumables for adequate hand-hygiene was limited (**Table 3**). Anal cleansing materials were identified at 18.9% of toilet visits, and despite 89.7% of households reporting they washed their hands after toileting, only 41.0% (n=120) households had a handwashing facility (HWF), 49.0% (n=166/339) of which had soap and 85.8% (n=349/408) of which had access to water upon visits. 4.3% (n=13) of households had adequate management of animal faeces and 8.0% (n=24) of households had adequate waste management of household rubbish.

**Table 3.**
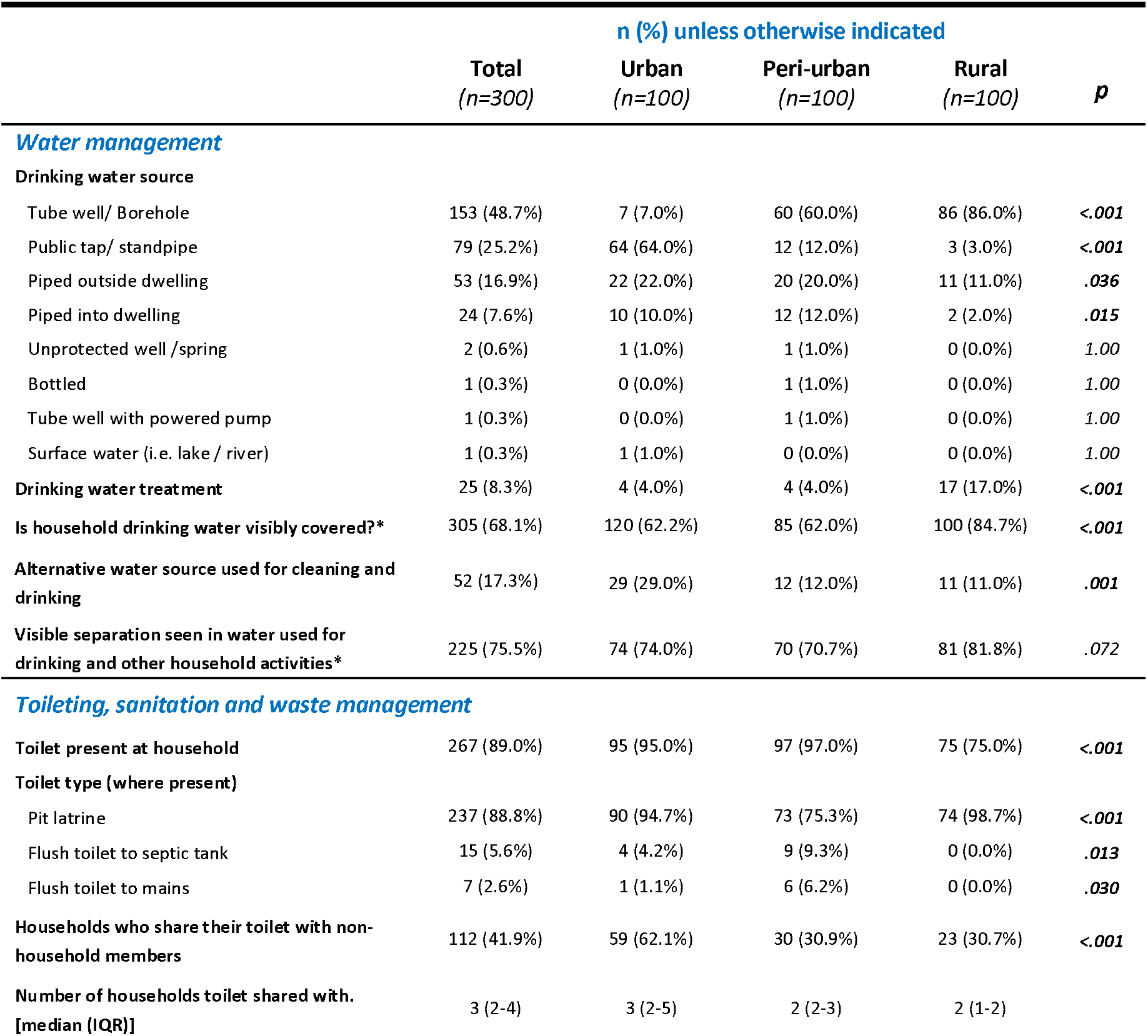
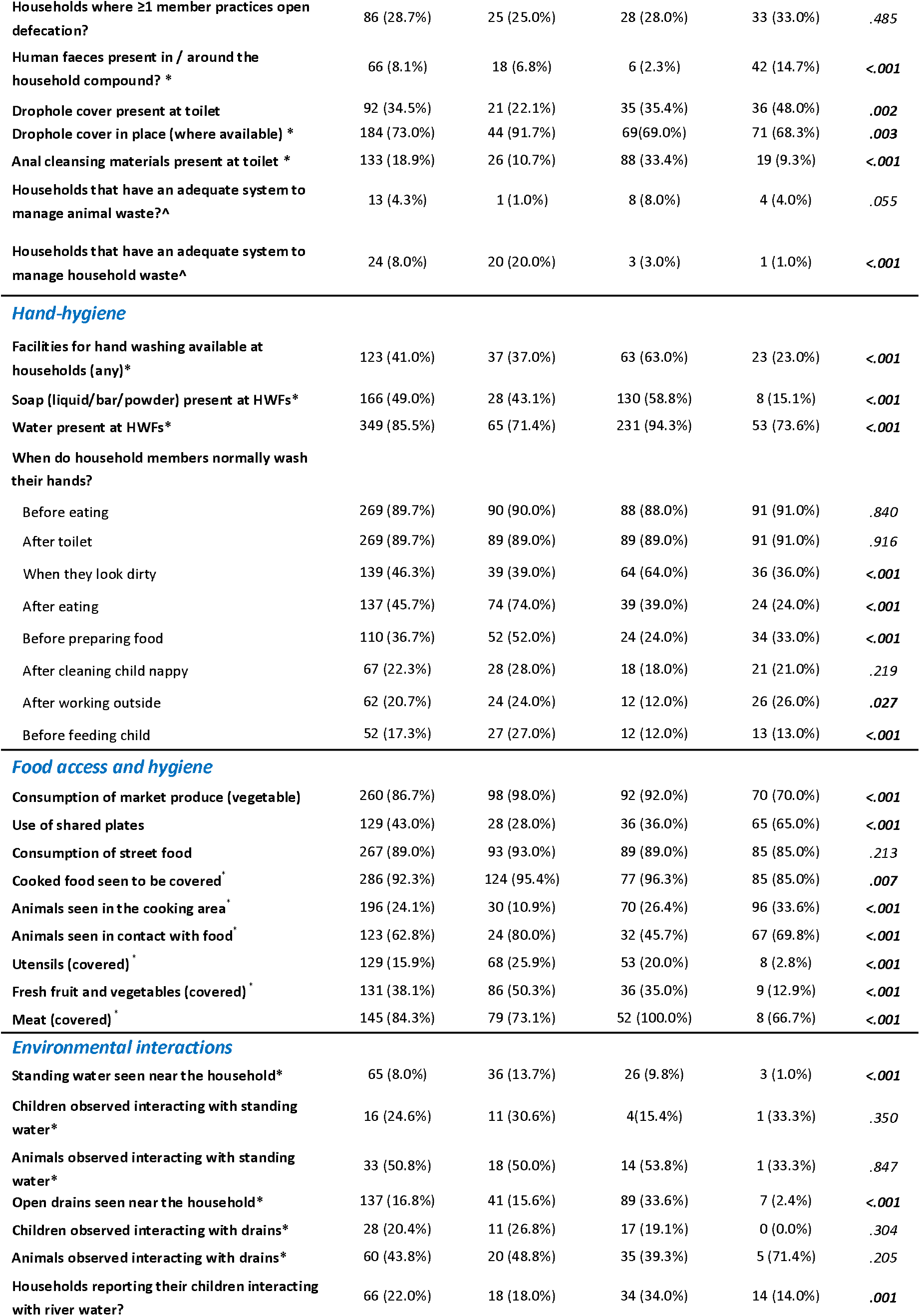
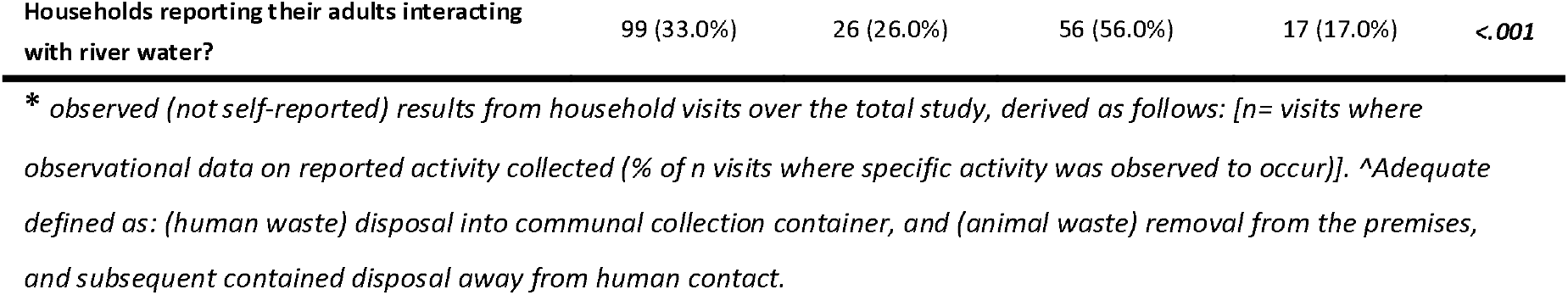
Baseline environmental health infrastructure, practices and environmental exposures

Households relied on local markets for purchasing vegetables (86.7%, n=260) and frequently ate street food (89.0% n=267). Cooked food was often seen to be covered (92.3%, n=286), but raw fruit and vegetables (38.1%, n=131), and cooking utensils (15.9% n=129) were often left uncovered. Where households owned animals, they were often seen in contact with human food (62.8% n=123) and were frequently present in food preparation areas (24.1% n=196).

Key environmental exposures included human contact, particularly children, with standing water, open drains and local rivers (**Table 3**). Observations of environmental sanitation practices were undertaken by study staff on each household visit, and where standing water was identified, children (24.6%, n=16) and animals (50.8%, n=33) were frequently observed interacting with it. High levels of interactions were also seen with the local sewerage system, with children and animals observed to be in contact with open drains 20.4% (n=28) and 43.8% (n=60) of the time respectively. Among the households, 22.0% (n=66) reported adults and 33.0% (n=99) reported children had regular contact with the local river network.

### Prevalence of ESBL E. coli and ESBL K. pneumoniae in humans, animals and the environment

In total 11,975 samples (2845 human stool, 973 animal stool and 8157 environmental samples) were cultured and ESBL *E. coli* or ESBL *K. pneumoniae* were isolated from 41.8% (n=1190) of human stool and 29.8% (n=290) of animal stool samples (**Fig 2 & S7 Table**). Animal species with particularly high rates of ESBL-E colonisation included pigs (56.8%, n=21/37) poultry (32.5%, n=148/455) and dogs (58.8% n= 30/51) (*Fig 3*). ESBL *E. coli* or ESBL *K. pneumoniae* were also isolated from a range of household environment, hand-hygiene, food, and community environment samples, with 66.2% (n=339/512) of river water samples and 46.0% (n=138/300) of drain samples having ESBL-E present.

**Figure 2.**
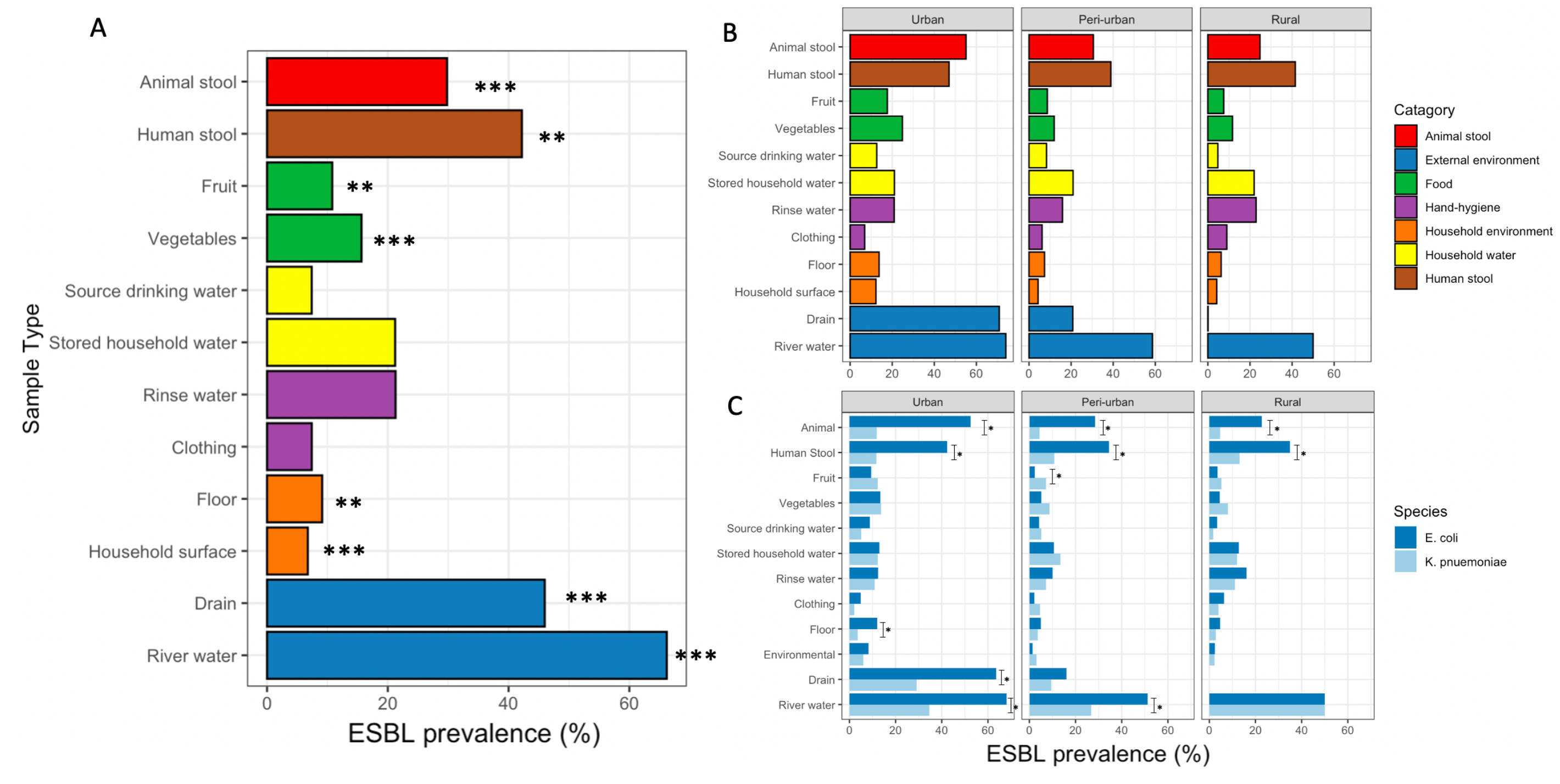
A) Proportion of samples positive for ESBL-E (*E. coli* ***<u>or</u>*** ESBL *K. pneumoniae*), at urban, peri-urban and rural households, coloured by category. Regional differences in the prevalence of ESBL for each sample types have been highlighted [*\*p >0*.*05, **p >0*.*01,and ***p >0*.*001*]. B) Breakdown of urban, peri-urban and rural rates of ESBL, coloured by sample category. C) Proportion of the household human stool, animal stool and environmental samples positive for ESBL *E. coli* or ESBL *K. pneumoniae*, stratified by sample type, bacterial species and region. Variations in the proportion of ESBL *E. coli* vs ESBL *K. pneumoniae* by sample type are highlighted by *(X^2^, p = <.001).

**Figure 3.**
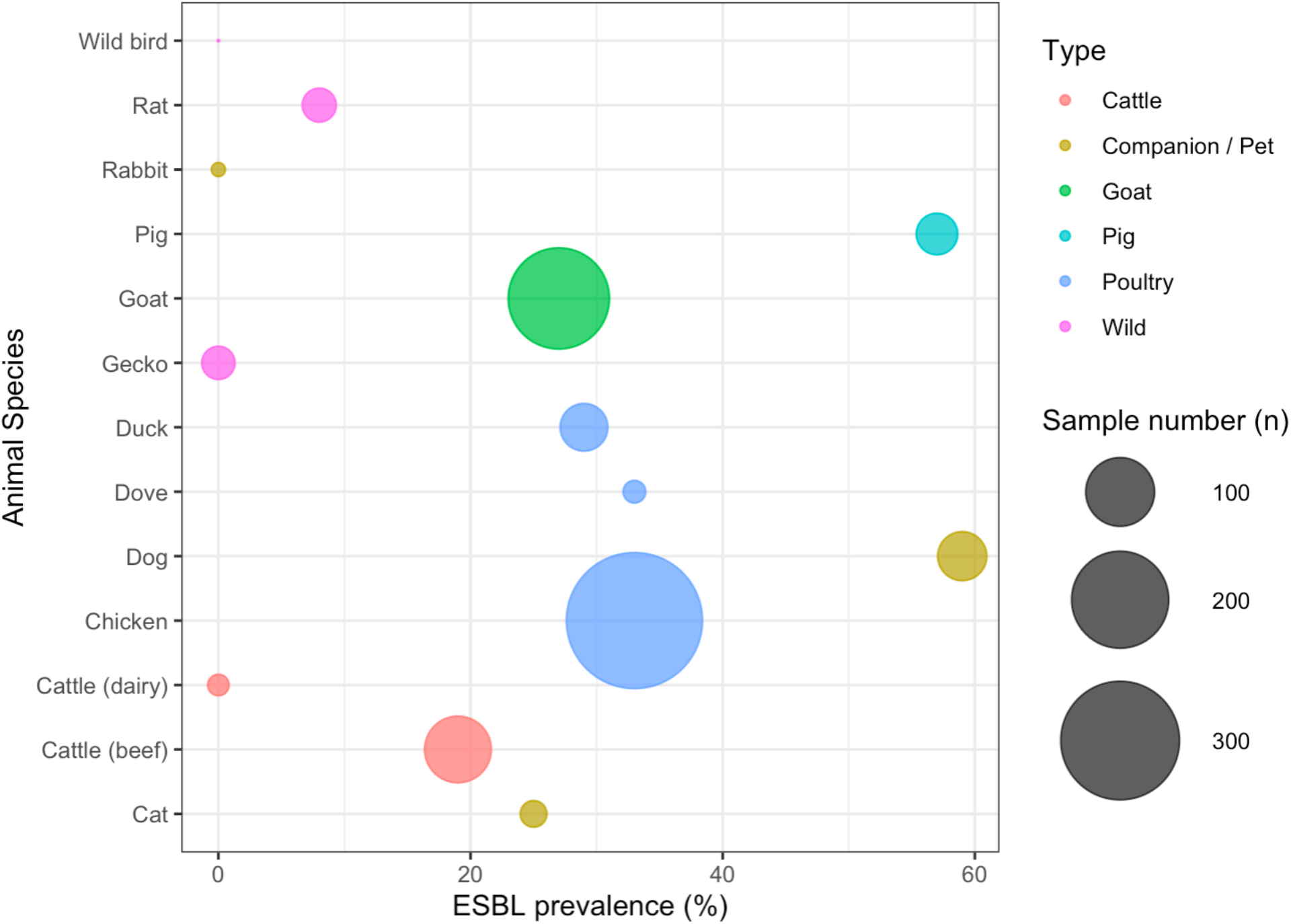
Bubble plot of ESBL-E (*E. coli* ***<u>or</u>*** *K. pneumoniae*) prevalence in animal stool samples, stratified by species, and coloured by animal type. The volume of the circle represents the number of samples processed for each species.

Amongst households with longitudinal follow-up 97.9% (n=191/195) had ≥1 ESBL colonised household member and 41.7% (n=50/120) of the households that owned animals had ≥1 ESBL colonised animal stool during the study period. 55.4% (n=108/195) of the households contained EBSL contaminated food and 45.6% (n=89/195) had contaminated environments at some point during the study. Longitudinal follow up revealed a high degree of flux (loss and acquisition) of ESBL-E in human household members, with 78.7% (n=588/747) of individuals undergoing longitudinal sampling having ESBL at any point (**S2a-c Figs**). Marked regional and seasonal regional differences in the prevalence of ESBL colonisation and contamination were noted (**Fig 2, Table 4**). Higher rates of ESBL-E were found in the urban settings, inclusive of animal stool, human stool, food, the household environment, and local drainage and river networks. Wet season was associated with a greater degree of ESBL presence in human stool, animal stool, stored drinking water and household floors and environments (**Table 4**).

**Table 4.** Seasonal variations in ESBL prevalence of household samples.

| Sample Type | ESBL prevalence by season (% , SD) |  | p |
| --- | --- | --- | --- |
|  | Wet season<br>(Nov-Apr) | Dry season<br>(May-Oct) |  |
| Human stool | 47.2% (49.9) | 36.6% (48.2) | <.001 |
| Animal stool | 33.3% (47.2) | 25.5% (43.6) | .010 |
| Food | 14.4% (35.1) | 12.3% (32.9) | .338 |
| Drinking water | 26.2% (44.0) | 15.2% (35.9) | <.001 |
| Source water | 6.5% (24.7) | 8.8% (28.3) | .413 |
| Household surfaces | 8.8% (28.3) | 4.5% (20.8) | <.001 |
| Household floor | 11.5% (31.9) | 6.6% (24.9) | .031 |
| Clothing | 9.1% (28.9) | 5.6% (23.0) | .087 |
| Hand-contact samples | 25.8% (43.9) | 17.9% (38.4) | .057 |
| Household drains | 44.7% (49.9) | 48.2% (50.2) | .648 |
| River water | 69.1% (46.3) | 62.9% (48.4) | .164 |

<sup>a</sup>p values generated by $\chi^2$ test

### One health factors associated with ESBL-E colonisation

PCA was used to describe regional differences and similarities in the individual-level, household-level, and environmental contamination variables (**Fig 4 & S3a-c Fig**). The first two individual-variable PCA coordinates explained 30.5% of the variation in the 12 included variables, defining orthogonal axes of age/employment and illness/ABU (**S3a Fig**). Household-variable PC1 and 2 explained 25.6% of variation in the 23 included variables, and highlighted that some animal and environmental health exposures tended to cluster together (**S3b Fig**). Environmental-contamination PC1 and 2 explained 38.7% of variation in the 9 environmental contamination variables and suggested that presence of ESBL-E in one household location was associated with ESBL-E in other household locations (**S3c Fig**).

**Figure 4.**
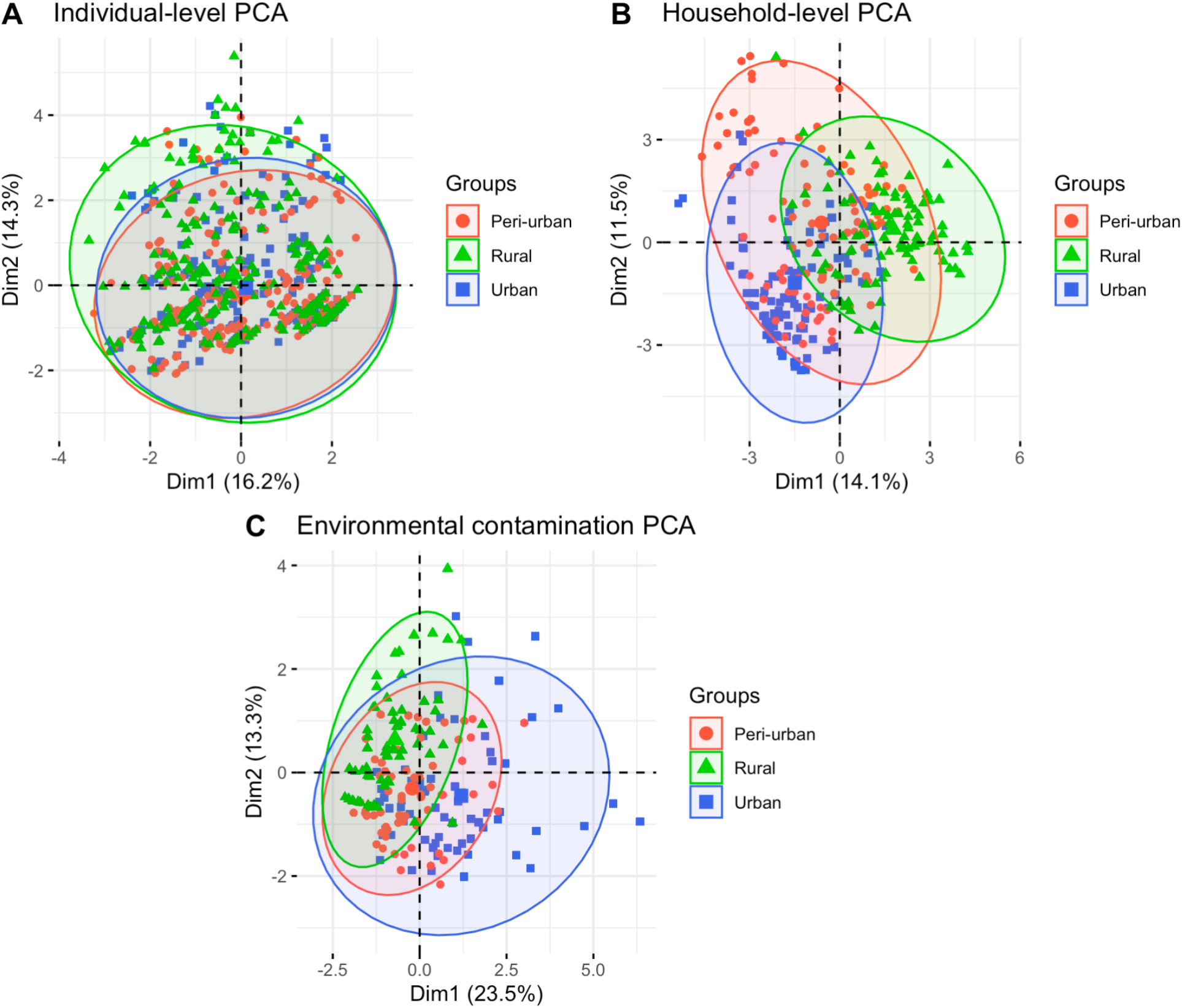
Confidence ellipses of regional effects exhibited by the (a) individual-level dataset, (b) household level dataset and (c) environmental contamination dataset, from the first 2 PCs. Points in 4A represent individuals but points in 4B and 4C represent households

Projection of individuals or households onto PCA (**Fig 4**) space stratified by region demonstrates that individuals are similar across regions (**Fig 4a**), but there are regional differences in distributions of household-level (**Fig 4b**) and environmental contamination (**Fig 4c**) variables, consistent with differences in animal husbandry and environmental health behaviours, and ESBL-E contamination across urban, peri-urban, and rural areas.

To identify regional differences in human ESBL-E colonisation, we constructed mixed effect logistic regression models. Variable selection resulted (**Tables S8a-b**) in 24 fixed effect predictor variables for ESBL *E. coli* and 14 fixed effect predictor variables for *K. pneumoniae*, as well as individual and household random effects. Here, the key risk associated with both human ESBL *E. coli* (*Fig 5*) and ESBL *K. pneumoniae* (**Fig 6**) colonisation was the presence of the wet season (*E. coli*: aOR = 1.66, 95%CrI: 1.38-2.00 / *K. pneumoniae*: aOR =2.12, 95%CrI: 1.63-2.76).

**Figure 5.**
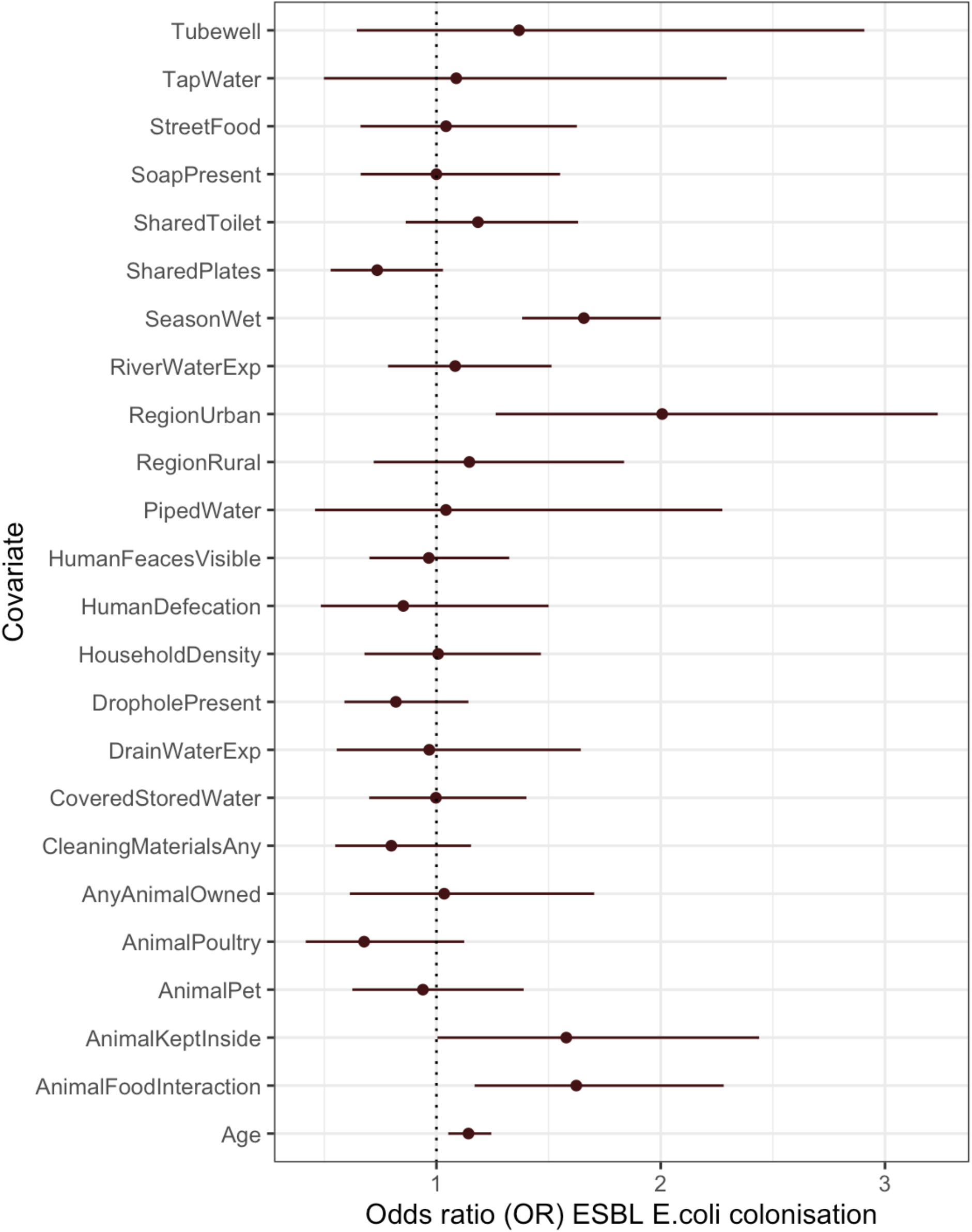
Parameter estimates for the fixed-effects used in a multivariable model of ESBL *E. coli* colonisation, expressed as odds ratios with 95% CrI. The distribution of random effects are visualised in **S4a Fig**.

**Figure 6.**
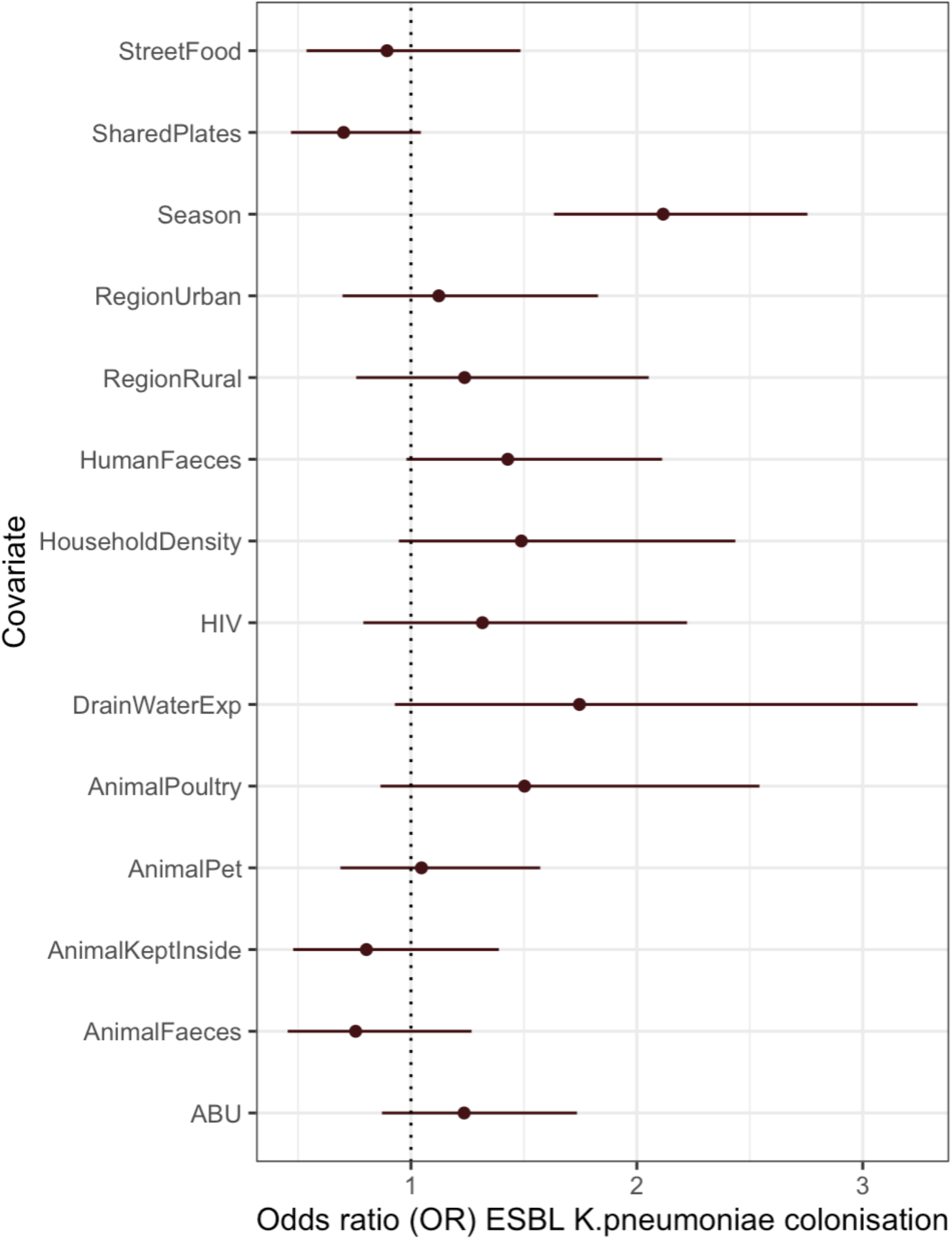
Parameter estimates for the fixed-effects used in a multivariable model of ESBL *K. pneumoniae* colonisation, expressed as odds ratios with 95% CrI. The distribution of random effects are visualised in **S4b Fig**.

Species-specific (i.e ESBL *E. coli* vs ESBL *K. pneumoniae*) risks other than the wet season were identified. Human ESBL *E. coli* colonisation was associated with advanced age (aOR = 1.14 per unit increase on log scale, 95%CrI: 1.05-1.25), within households where animals were kept inside the house (aOR = 1.58, 95%CrI: 1.00-2.43) or in those households where animals were observed interacting with food (aOR = 1.62, 95%CrI: 1.17-2.28) (*Fig 5*). Accounting for these factors did not fully explain the increased urban EBSL *E. coli* prevalence, therefore living in the urban environment was shown to be a risk compared to the peri-urban site (aOR = 2.01, 95%CrI: 1.26-3.24).

Human ESBL *K. pneumoniae* colonisation was only shown to be associated with the wet season (aOR = 2.12, 95%CrI: 1.63-2.76), and there was no effect of urbanisation seen. However, there was a trend towards increased risk seen in households with sub-optimal environmental sanitation, including those where human faecal contamination was observed in the internal/external environment (aOR = 1.43, 95%CrI: 0.98-2.11), those where household members interacted with drain water (aOR = 1.75, 95%CrI: 0.93-3.24) and households with increasing density of members (aOR = 1.49, 95%CrI: 0.85-2.60) (**Fig 6**). Antibiotic usage was not identified as a risk for either ESBL *E. coli* or ESBL *K. pneumoniae* colonisation.

To explore the possibility that the difference in ESBL prevalence between urban, peri-urban and rural households could be explained by covariates exerting a different effect in different regions we fit models with a covariate by region interaction (**S1a-b Figs & S9a-b Tables**). The only covariate that seemed to exert a different effect across regions was seasonality: ESBL *E. coli* colonisation was associated with a higher risk in the peri-urban site during the wet season (aOR = 2.66, 95%CrI: 1.90-3.69) compared to the urban (aOR = 1.38, 95%CrI: 1.01-1.90) or rural (aOR = 1.25, 95%CrI: 0.90-1.72) site. However, confidence intervals were wide making it difficult to draw further conclusions, and no other differential regional effects were identified for ESBL *E. coli* or ESBL *K. pneumoniae*.

## Discussion

We have taken a One Health approach in this large, multi-site, longitudinal cohort study of ESBL-E in households in three regions with different degrees of urbanisation in southern Malawi. We identified extremely high levels of ESBL-E colonisation in humans and animals alongside extensive ESBL contamination of the environments in all urban, peri-urban and rural communities studied. We describe paucity of household environmental health infrastructure and access to materials that promote safe toileting, adequate sanitation, effective hand-hygiene or acceptable waste management and also note the variations in environmental practice and infrastructure between sites, with regional differences in ESBL-E colonisation rates and levels of environmental contamination. In particular, we highlight the effects of urbanisation and seasonal rainfall on ESBL colonisation, with the highest rates of human and animal ESBL-E colonisation identified in the urban site compared to other regions and a higher prevalence of ESBL-E colonisation and contamination of household surfaces and drinking water noted in the wet season.

ABU is increasing globally, particularly within the commercial livestock production sector, and this has been highlighted by the many authorities as an important driver of AMR (13,14). Moderate levels of human ABU were reported, with under 5s and inhabitants in rural communities having a higher ABU overall. Antibiotic prescription was limited to oral co-trimoxazole, amoxicillin or metronidazole. These medications are on the international essential medicine lists for treatment of a number of infectious conditions (12–14), and the use of only a small number of antibiotics is consistent with findings from previous local descriptions from community-focussed ABU research (15). ABU in household animals was limited in our study, despite reports of animal illness. However, we did not include households practicing intensive small-scale farming, and in this context animals are regularly administered antimicrobials (15), kept in cramped conditions, and have a high rate of AMR colonisation, including ESBL *E. coli* (16–18).

Despite low rates of reported ABU, we observed high rates of ESBL-E colonisation in animals, especially in the urban region, consistent with evidence from other LMIC settings where sharing of ESBL-producing bacteria between household members and domestic animals / livestock (19,20) have been reported (21,22). Local animal husbandry practices including the proximity and location of animal co-habitation (23–25), household attitudes to animal and human waste management in the shared environment and animal interactions with key external environments are likely to promote ESBL transmission. In this study, we found that animals were regularly in contact with heavily contaminated external environments (open drains and rivers) and also with food / food preparation areas within the household, which in turn were associated with higher odds of ESBL-E colonisation in humans (aOR = 1.62, 95%CrI: 1.17-2.28). In addition to this, animals, especially poultry, were frequently kept inside the household providing an increased risk to residents (aOR = 1.58, 95%CrI: 1.00-2.43) and animal and human waste was commonly identified in or around the household. The dynamics of these animal-human-waste interactions may drive the maintenance and transmission for ESBL-E within animals, especially in the urban setting.

We found a paucity of environmental health infrastructure, a scarcity of access to preventative hygiene materials, and high-risk behavioural proxies for faecal-oral acquisition at a household level. This included frequent interactions with open drains and rivers within the local vicinity, where there are consistently high levels of ESBL bacteria, likely derived from inadequate human and animal waste management. Our data therefore point to unrestricted shedding of human and animal waste into an unprotected environment as playing a key role in AMR transmission, whether acquisition is from household members (i.e. human-human transmission), co-located animals (i.e. human-animal transmission or vice versa) or transmission to and from the external environment. We propose that availability of environmental health infrastructure and services, hygiene practices and environmental hygiene govern the transmission of ESBL bacteria in Malawian communities. Further research into the protective effects of environmental health interventions should be considered to determine their impact on the transmission and prevalence of ESBL colonisation within and between the three One-Health sectors.

The effect of urbanisation, where there was high prevalence of ESBL contamination of food, household surfaces, floors and the external environment, was of particular importance. This was likely to have been heavily influenced by a combination of animal and environmental health factors, including the ability of the local environment and environmental hygiene infrastructure to cope with extreme weather events such as high rainfall. Future interventions and policy designed to interrupt AMR transmission should be cognisant of regional differences in AMR-prevalence, there will not be a “one-size-fits-all” solution to community transmission of AMR.

There are limitations to our study. Due to the study design, the majority of our demographic, ABU, animal husbandry data and a portion of our environmental health data was obtained from self-reported responses, which are subject to recall bias. Self-reported hygiene behaviour data can especially be prone to recall bias, especially in our setting (26). To mitigate this, where possible we used observed data from checklists in place of self-reported data. Further the AMR data presented are solely phenotypic and future whole genome sequencing of the archive from this study is underway and will permit transmission modelling to be undertaken.

In conclusion, we found a staggeringly high prevalence of ESBL-E colonisation in humans and animals, together with extensive ESBL-E contamination of the households and broader environment (i.e. rivers and drains) in southern Malawi. The findings also highlight the key role that environmental health infrastructure and behavioural proxies have on driving human community carriage of ESBL bacteria in southern Malawi. We therefore propose that without adequate efforts to reduce ESBL contamination of the shared environment, both at a household level and community level, we are unlikely to control ESBL transmission in this setting. Lastly, regional differences in AMR-prevalence exist, which are influenced by regionally-specific environmental health and animal husbandry factors. Therefore, future interventions aimed at interrupting ESBL-E transmission should be sensitive to regional differences and tailored accordingly. These findings form a starting point from which a more detailed understanding of the drivers and ecological niches for ESBL *E. coli* and ESBL *K. pneumoniae* AMR in Malawi, and comparable LMIC settings can be built.

## Supporting information

Supplementary household data

Supplementary WASH data

Supplementary participant data

Supplementary laboratory data

## Data Availability

All data produced in the present work are contained in the manuscript

## Acknowledgements

We would like to thank the local communities for their acceptance of this study, and the households and participants who took part. We would also like to thank the wider DRUM consortium (https://www.drumconsortium.org/about/the-drum-team) for their advice, guidance and support.

## Supplementary data

- **S1a Table**: Individual-level variables selected from the CRFs
- **S1b Table**: Household, WASH and sampling variables selected from the CRFs
- **S1c Table**: Outcome variables and covariates.
- **S2 Table**: STROBE checklist.
- **S3a Table**: Household data.
- **S3b Table**: WASH data.
- **S3c Table**: Individual data
- **S3d Table**: Laboratory data
- **S4a Table:** Baseline household ABU from urban, peri-urban and rural sites.
- **S4b Table**: AMU in different age groups
- **S5 Table**: Domestic animal and livestock ownership and husbandry
- **S6 Table**: Healthcare choices for household animals
- **S7 Table**: Numbers of samples screened for ESBL *E. coli* and ESBL *K. pneumoniae*, stratified by sample type and region.
- **S8a Table**: Regional univariable analysis of WASH and individual variables against human ESBL *E. coli* colonisation
- **S8b Table**: Regional univariable analysis of WASH and individual variables against human ESBL *K. pneumoniae* colonisation
- ***S9a Table***: Table of parameter testing for regional adjustment of variables included in the ESBL *E. coli* mixed effects model
- **S9b Table**: Table of parameter testing for regional adjustment of variables included in the ESBL *K. pneumoniae* mixed effects model
- **S1a Fig**: Parameter estimates for the fixed-effects used in a multivariable model of ESBL *E. coli*
- **S1b Fig**: Parameter estimates for the fixed-effects used in a multivariable model of ESBL *K. pneumoniae* colonisation
- **S2a Fig:** Facet Plot showing flux of human ESBL-E colonisation amongst *urban* households
- **S2b Fig**: Facet Plot showing flux of human ESBL-E colonisation amongst *per-urban* households
- **S2c Fig:** Facet Plot showing flux of human ESBL-E colonisation amongst *rural* households
- **S3a Fig:** PCA analysis of individual variables
- **S3b Fig:** PCA analysis of household variables
- **S3c Fig:** PCA analysis of environmental contamination variables
- **S4 Fig:** Random effects from Bayesian multivariable models of (a) ESBL *E. coli*, and (b) ESBL *K. pneumoniae*.

