## Supplementary data tables and figures for "Investigating risks for human colonisation with extended spectrum beta-lactamase producing *E. coli* and *K. pneumoniae* in Malawian households: a one health longitudinal cohort study"

**S1a Table.** Individual-level variables selected from the CRFs for analysis, including any groupings, outputs and transformations undertaken.

| Variable name | Description |
|---|---|
| Age* | Continuous, age at enrolment (years) |
| Male | Binary, {1 = male, 0 = female} |
| Religion (Christianity) | Binary, {1 = faith reported as Christianity, 0 = other religion practiced} |
| School (attendance) | Binary, {1 = attend school, 0 = do not attend school} |
| School (work) | Binary, {1 = work in/at school, 0 = do not work in/at school} |
| Healthcare (work) | Binary, {1 = work in/at hospital, 0 = do not work in/at hospital} |
| Hospital admission | Binary, {1 = admitted overnight to hospital in last 6 months, 0 = not admitted over last 6 months} |
| Hospital guardian | Binary, {1 = been a guardian at hospital in last 6 months, 0 = not been a guardian at hospital in last 6 months) |
| Employed | Binary, {1 = have regular job at recruitment, 0 = no job at recruitment} |
| Residency | Binary, {1 = resident for year or more at household, 0 = not resident for year} |
| Travel (Outside region) | Binary, {1 = travel outside region (any purpose in last 6 months, 0 = no travel outside area in last 6 months} |
| HIV status | Binary, {1 = HIV reactive, 0 = HIV non-reactive or unknown} |
| TB history | Binary, {1 = ever had diagnosis of TB, 0 = never had TB} |
| Comorbidities | Binary, {1 = 1 or more comorbidities, 0 = no comorbidities} |
| Medication | Binary, {1 = take any prescribed regular medication, 0 = do not take any regular medication} |
| Unwell 4 weeks | Binary, {1 = 1 or more unwell episodes in last 4 weeks, 0 = no illness episodes in last 4 weeks} |
| Unwell 3 months | Binary, {1 = 1 or more unwell episodes in last 3 months, 0 = no illness episodes in last 3 months} |
| ABU | Binary, {1 = 1 or more antibiotic courses taken in the last 6 months, 0 = no antibiotics taken in last 6 months} |

**log-transformed*

**S1b Table.** Household, WASH and sampling variables selected from the CRFs for analysis, including any outputs and transformations undertaken. Variables are grouped into reported, observed and laboratory categories and stratified by factor type.

| | | Variable | Description |
|---|---|---|---|
| Household Factors | Reported | Number of people living in house* | Continuous, number of people cohabiting at baseline. |
| Household Factors | Reported | Household income* | Continuous, household income (MK) at baseline. |
| Household Factors | Lab | Share household with ESBL colonised humans | Binary, {1 = yes [share with 1 or more ESBL colonised individuals within the same household], 0 = no [Do not share with ESBL colonised household members ]} |
| Sanitation Factors | Observed | Presence of drop hole cover | Binary, {1 = drop hole cover present, 0 = drop hole cover absent} |
| Sanitation Factors | Observed | Cleansing materials at toilet | Binary, {1 = cleansing material [any type] present, 0 = cleansing material [any type] absent} |
| Sanitation Factors | Observed | Visible human defecation | Binary, {1 = visible human stool [adult or child], 0 = no visible human stool} |
| Sanitation Factors | Reported | Use of pit latrine | Binary, {1 = use pit latrine, 0 = use other toilet type, or do not have toilet} |
| Sanitation Factors | Reported | Toilet presence (any) at household | Binary, {1 = toilet present, 0 = toilet absent} |
| Sanitation Factors | Reported | Open human defecation | Binary, {1 = open defecation reported [by 1 or more household members], 0 = no open defecation reported [by all household member]} |
| Sanitation Factors | Reported | Sharing household toilet with non-household members | Binary, {1 = shared toilet used, 0 = do not share their toilet external to the household} |
| Sanitation Factors | Reported | Absence of disposal mechanism for animal waste | Binary, {1 = no disposal mechanism for animal faeces, 0 = dispose of animal faeces by either sweeping them away, putting into a refuse pit or re-using as manure} |
| Sanitation Factors | Lab | Household environmental ESBL contamination | Binary, {1 = yes [at any point in the household during study], 0 = no [at all points during study]} |
| Hygiene factors | Reported | Facilities for hand washing (all areas) at household | Binary, {1 = yes [present at one or more place within the household], 0 = no [present at no places within the household]} |
| Hygiene factors | Observed | Presence of soap at (any) HWF | Binary, {1 = yes [present at one or more HWFs within the household], 0 = no [present at no HWFs within the household]} |
| Hygiene factors | Lab | Household rinse water ESBL contamination | Binary, {1 = yes [at any point in the household during study], 0 = no [at all points during study]} |
| Food Factors | Reported | Eat street food | Binary, {1 = yes [supplement diet with street food at some points], 0 = no [never buy street food]} |
| Food Factors | Reported | Eat from shared plates | Binary, {1 = yes [use shared plates], 0 = no [do not use shared plates]} |
| Food Factors | Reported | Buy vegetables or fruit from the market | Binary, {1 = yes [use vegetables or fruit from the market on any occasion], 0 = no [do not use market food or fruit]} |

| | | | |
|---|---|---|---|
| | Lab | Household food ESBL contamination | Binary, {1 = yes [at any point in the household during study], 0 = no [at all points during study]} |
| Water Factors | Observed | Is water stored in the house covered? | Binary, {1 = yes [water stored at the house covered], 0 = no [water not stored at the house covered]} |
| | Reported | Is water stored in the house? | Binary, {1 = yes [water stored at the house], 0 = no [water not stored at the house]} |
| | | Drinking water source piped into household | Binary, {1 = yes [water source from outside the household], 0 = no [water source from pipe inside or directly outside the household]} |
| | | Drinking water source kiosk | Binary, {1 = yes [water source from outside the household], 0 = no [water source from pipe inside or directly outside the household]} |
| | | Drinking water source tubewell | Binary, {1 = yes [water source from outside the household], 0 = no [water source from pipe inside or directly outside the household]} |
| | | Alternative water used for cleaning utensils | Binary, {1 = yes [different water used for cleaning utensils than for drinking], 0 = no [same water used for cleaning utensils as drinking]} |
| | Lab | Household source water ESBL contamination | Binary, {1 = yes [at any point in the household during study], 0 = no [at all points during study]} |
| | | Household stored water ESBL contamination | Binary, {1 = yes [at any point in the household during study], 0 = no [at all points during study]} |
| Animal Factors | Observed | Animal faeces seen around the area | Binary, {1 = yes [any animal faeces seen around the household at any point], 0 = no [no animal faeces ever seen around the household]} |
| | | Evidence of animal contact with food | Binary, {1 = yes [animal seen in contact with food], 0 = no [no animal seen in contact with food]} |
| | Reported | Does the household own any animals? | Binary, {1 = yes [household owns 1 or more animals], 0 = no [no animals owned by household]} |
| | | Own cattle or ruminant | Binary, {1 = yes [household owns 1 or more animals], 0 = no [no animals owned by household]} |
| | | Own poultry | Binary, {1 = yes [household owns 1 or more animals], 0 = no [no animals owned by household]} |
| | | Own pet / companion animal | Binary, {1 = yes [household owns 1 or more animals], 0 = no [no animals owned by household]} |
| | | Own pigs | Binary, {1 = yes [household owns 1 or more animals], 0 = no [no animals owned by household]} |
| | | Animals (any species) kept inside the house? | Binary, {1 = yes [if animals owned - they kept inside the house], 0 = no [if animals owned - they are not kept inside the house]} |
| | Lab | Household animal ESBL contamination | Binary, {1 = yes [at any point in the household animals during study], 0 = no [at all points during study]} |
| Broader Environme | Observed | Accumulation of water / wastewater (household environment) | Binary, {1 = yes [water seen external to the household], 0 = no [water not seen external to the household]} |

| | | | |
|---|---|---|---|
| | **Reported** | Household member interaction with river water | Binary, {1 = yes [any adult or child at the household reportedly interact with river water], 0 = no [no adult or child at the household even interact with river water]} |
| | | Household member interaction with drains | Binary, {1 = yes [any adult or child at the household reportedly interact with drains], 0 = no [no adult or child at the household even interact with drains]} |
| | **Lab** | Drain ESBL contamination | Binary, {1 = yes [at any point during study], 0 = no [at all points during study]} |
| | | River ESBL contamination | Binary, {1 = yes [at any point during study], 0 = no [at all points during study]} |

**log-transformed*

**S1c Table.** Outcome variables and covariates.

| Dependant variable | Description |
|---|---|
| ESBL (positive) | Binary, {1 = ESBL positive at single episode [with either KPN or EC], 0 = ESBL negative at single episode [with either KPN or EC]} |
| ESBL-E (positive) | Binary, {1 = ESBL *E. coli* positive at single episode, 0 = ESBL *E. coli* negative at single episode} |
| ESBL-K (positive) | Binary, {1 = ESBL *K. pneumoniae* positive at single episode, 0 = ESBL *K. pneumoniae* negative at single episode} |
| **Covariates** | |
| Region | Categorical, {Urban / Peri-urban / rural} |
| Season | Binary, {1 = wet (October-April), 0 = dry (May-September)} |

**S2 Table:** STROBE Statement

| | Item No | Recommendation | Page No |
|---|---|---|---|
| **Title and abstract** | 1 | (*a*) Indicate the study's design with a commonly used term in the title or the abstract | 1 (title) |
| | | (*b*) Provide in the abstract an informative and balanced summary of what was done and what was found | 3 |
| **Introduction** | | | |
| Background/rationale | 2 | Explain the scientific background and rationale for the investigation being reported | 5-6 |
| Objectives | 3 | State specific objectives, including any prespecified hypotheses | 6 |
| **Methods** | | | |
| Study design | 4 | Present key elements of study design early in the paper | 6-7, including reference to online protocol |
| Setting | 5 | Describe the setting, locations, and relevant dates, including periods of recruitment, exposure, follow-up, and data collection | 6, setting and location described in protocol paper which is referenced in the text. All other aspects completed |
| Participants | 6 | (*a*) *Cohort study*—Give the eligibility criteria, and the sources and methods of selection of participants. Describe methods of follow-up<br>*Case-control study*—Give the eligibility criteria, and the sources and methods of case ascertainment and control selection. Give the rationale for the choice of cases and controls<br>*Cross-sectional study*—Give the eligibility criteria, and the sources and methods of selection of participants | 6 |
| | | (*b*) *Cohort study*—For matched studies, give matching criteria and number of exposed and unexposed<br>*Case-control study*—For matched studies, give matching criteria and the number of controls per case | NA |
| Variables | 7 | Clearly define all outcomes, exposures, predictors, potential confounders, and effect modifiers. Give diagnostic criteria, if applicable | 7-9 and S1a-c Tables. |
| Data sources/ measurement | 8* | For each variable of interest, give sources of data and details of methods of assessment (measurement). Describe comparability of assessment methods if there is more than one group | S1a-c Tables. Statistical methods explained in pages 7-8. Data included in S3a-e |
| Bias | 9 | Describe any efforts to address potential sources of bias | 6, 8-9 and 22. |
| Study size | 10 | Explain how the study size was arrived at | 6, and detailed in protocol paper referenced in the manuscript. |

| | | | |
|---|---|---|---|
| Quantitative variables | 11 | Explain how quantitative variables were handled in the analyses. If applicable, describe which groupings were chosen and why | S1a-c Tables. Statistical methods explained in pages 7-8. |
| Statistical methods | 12 | (*a*) Describe all statistical methods, including those used to control for confounding | 7-8 |
| | | (*b*) Describe any methods used to examine subgroups and interactions | 7-8 and S1a-b figs |
| | | (*c*) Explain how missing data were addressed | 8 |
| | | (*d*) *Cohort study*—If applicable, explain how loss to follow-up was addressed<br>*Case-control study*—If applicable, explain how matching of cases and controls was addressed<br>*Cross-sectional study*—If applicable, describe analytical methods taking account of sampling strategy | 8 |
| | | (*e*) Describe any sensitivity analyses | NA |

Continued on next page

| | | | |
|---|---|---|---|
| **Results** | | | |
| Participants | 13* | (a) Report numbers of individuals at each stage of study—eg numbers potentially eligible, examined for eligibility, confirmed eligible, included in the study, completing follow-up, and analysed | 11 |
| | | (b) Give reasons for non-participation at each stage | 11 |
| | | (c) Consider use of a flow diagram | 11 |
| Descriptive data | 14* | (a) Give characteristics of study participants (eg demographic, clinical, social) and information on exposures and potential confounders | 11-12 |
| | | (b) Indicate number of participants with missing data for each variable of interest | NA |
| | | (c) *Cohort study*—Summarise follow-up time (eg, average and total amount) | 9 |
| Outcome data | 15* | *Cohort study*—Report numbers of outcome events or summary measures over time | 17-20 |
| | | *Case-control study*—Report numbers in each exposure category, or summary measures of exposure | NA |
| | | *Cross-sectional study*—Report numbers of outcome events or summary measures | NA |
| Main results | 16 | (*a*) Give unadjusted estimates and, if applicable, confounder-adjusted estimates and their precision (eg, 95% confidence interval). Make clear which confounders were adjusted for and why they were included | 17-20 |
| | | (*b*) Report category boundaries when continuous variables were categorized | 10-11, 14-15, |
| | | (*c*) If relevant, consider translating estimates of relative risk into absolute risk for a meaningful time period | NA |
| Other analyses | 17 | Report other analyses done—eg analyses of subgroups and interactions, and sensitivity analyses | Supplementary data |
| **Discussion** | | | |
| Key results | 18 | Summarise key results with reference to study objectives | 20 |
| Limitations | 19 | Discuss limitations of the study, taking into account sources of potential bias or imprecision. Discuss both direction and magnitude of any potential bias | 22 |
| Interpretation | 20 | Give a cautious overall interpretation of results considering objectives, limitations, multiplicity of analyses, results from similar studies, and other relevant evidence | 22-23 |
| Generalisability | 21 | Discuss the generalisability (external validity) of the study results | 22-23 |
| **Other information** | | | |
| Funding | 22 | Give the source of funding and the role of the funders for the present study and, if applicable, for the original study on which the present article is based | Listed during submission |

**S4a Table.** Household (baseline) ABU from urban, peri-urban and rural sites.

| Variable | Site | Total antibiotics | Amoxicillin | Benzylpenicillin | Cefuroxime | Cefixime | Ciprofloxacin | Co-trimoxazole | Doxycycline | Erythromycin | Gentamicin | Metronidazole | Other or Unknown |
|---|---|---|---|---|---|---|---|---|---|---|---|---|---|
| | | **Antibiotic choice^** | | | | | | | | | | | |
| **Antibiotic usage in household participants** | Urban | n=63 | n=24 (38.1%) | n=2 (3.2%) | n=0 (0.0%) | n=0 (0.0%) | n=2 (3.2%) | n=20 (31.7%) | n=0 (0.0%) | n=2 (3.2%) | n=3 (4.8%) | n=7 (11.1%) | n=3 (4.8%) |
| | Peri-urban | n=40 | n=9 (22.5%) | n=0 (0.0%) | n=0 (0.0%) | n=0 (0.0%) | n=3 (7.5%) | n=17 (42.5%) | n=0 (0.0%) | n=1 (2.5%) | n=1 (2.5%) | n=8 (20.0%) | n=1 (2.5%) |
| | Rural | n=78 | n=31 (39.7%) | n=1 (1.3%) | n=1 (1.3%) | n=1 (1.3%) | n=2 (2.6%) | n=28 (35.9%) | n=2 (2.6%) | n=1 (1.3%) | n=2 (2.6%) | n=8 (10.3%) | n=1 (1.3%) |
| **Antibiotic used in last 4 weeks** | Urban | n=21 | n=5 (23.8%) | n=0 (0.0%) | n=0 (0.0%) | n=0 (0.0%) | n=1 (4.8%) | n=10 (47.6%) | n=0 (0.0%) | n=0 (0.0%) | n=1 (4.8%) | n=2 (9.5%) | n=2 (9.5%) |
| | Peri-urban | n=24 | n=4 (16.7%) | n=0 (0.0%) | n=0 (0.0%) | n=0 (0.0%) | n=1 (4.2%) | n=12 (50.0%) | n=0 (0.0%) | n=0 (0.0%) | n=1 (4.2%) | n=5 (20.8%) | n=1 (4.2%) |
| | Rural | n=29 | n=12 (41.4%) | n=0 (0.0%) | n=0 (0.0%) | n=0 (0.0%) | n=1 (3.4%) | n=12 (41.4%) | n=0 (0.0%) | n=0 (0.0%) | n=0 (0.0%) | n=3 (10.3%) | n=1 (3.4%) |
| **Antibiotic used in last 4 weeks to 3 months** | Urban | n=24 | n=8 (33.3%) | n=1 (4.2%) | n=0 (0.0%) | n=0 (0.0%) | n=1 (4.2%) | n=6 (25.0%) | n=0 (0.0%) | n=2 (8.3%) | n=1 (4.2%) | n=4 (16.7%) | n=1 (4.2%) |
| | Peri-urban | n=8 | n=3 (32.5%) | n=0 (0.0%) | n=0 (0.0%) | n=0 (0.0%) | n=1 (12.5%) | n=2 (25.0%) | n=0 (0.0%) | n=1 (12.5%) | n=0 (0.0%) | n=1 (12.5%) | n=0 (0.0%) |
| | Rural | n=33 | n=12 (36.4%) | n=1 (3.0%) | n=1 (3.0%) | n=0 (0.0%) | n=1 (3.0%) | n=11 (33.3%) | n=2 (6.0%) | n=0 (0.0%) | n=2 (6.0%) | n=3 (9.1%) | n=0 (0.0%) |
| **Antibiotic used in last 6 months* (healthcare)** | Urban | n=15 | n=9 (60.0%) | n=1 (6.7%) | n=0 (0.0%) | n=0 (0.0%) | n=0 (0.0%) | n=3 (20.0%) | n=0 (0.0%) | n=0 (0.0%) | n=1 (6.7%) | n=1 (6.7%) | n=0 (0.0%) |
| | Peri-urban | n=3 | n=0 (0.0%) | n=0 (0.0%) | n=0 (0.0%) | n=0 (0.0%) | n=1 (33.3%) | n=1 (33.3%) | n=0 (0.0%) | n=0 (0.0%) | n=0 (0.0%) | n=1 (33.3%) | n=0 (0.0%) |
| | Rural | n=15 | n=7 (46.7%) | n=0 (0.0%) | n=0 (0.0%) | n=1 (6.7%) | n=0 (0.0%) | n=4 (26.7%) | n=0 (0.0%) | n=1 (6.7%) | n=0 (0.0%) | n=2 (13.3%) | n=0 (0.0%) |
| **Antibiotic used at baseline** | Urban | n=3 | n=2 (66.7%) | n=0 (0.0%) | n=0 (0.0%) | n=0 (0.0%) | n=0 (0.0%) | n=1 (33.3%) | n=0 (0.0%) | n=0 (0.0%) | n=0 (0.0%) | n=0 (0.0%) | n=0 (0.0%) |
| | Peri-urban | n=5 | n=2 (40.0%) | n=0 (0.0%) | n=0 (0.0%) | n=0 (0.0%) | n=0 (0.0%) | n=2 (40.0%) | n=0 (0.0%) | n=0 (0.0%) | n=0 (0.0%) | n=1 (20.0%) | n=0 (0.0%) |
| | Rural | n=1 | n=0 (0.0%) | n=0 (0.0%) | n=0 (0.0%) | n=0 (0.0%) | n=0 (0.0%) | n=1 (100%) | n=0 (0.0%) | n=0 (0.0%) | n=0 (0.0%) | n=0 (0.0%) | n=0 (0.0%) |

*^Grey = Total usage, where antibiotics were selected by ≥1 households in region. Blue = cumulative total of antibiotics used. Yellow = antibiotic selected by ≥1 households in region. White = antibiotic not selected.*Provided to participant at healthcare services during an acute presentation.*

**S4b Table.** AMU in different age groups

| Reported ABU | | n (%) | | | |
|---|---|---|---|---|---|
| **Antibiotic use by age group**[$] | | **Child** | **Adolescent** | **Adult** | *p* |
| Antibiotic usage (total all regions) | NA | 49 (32.5%) | 39 (11.9%) | 59 (12.2%) | ***>.001*** |

*[$]Total Adult (>17) = 485, Adolescents (5-17) = 329, and Children <5 = 151*

**S5 Table.** Domestic animal and livestock ownership and husbandry

| **Household ownership** | n | | | |
|---|---|---|---|---|
| | **Total** | **Urban** | **Peri-urban** | **Rural** |
| **Total number of animals owned** | 2169 | 213 | 704 | 1252 |
| **Species of animals owned^** | | | | |
| Number of chickens | 919 | 152 | 315 | 452 |
| Number of doves | 442 | 10 | 250 | 182 |
| Number of ducks | 67 | 14 | 8 | 45 |
| Number of guinea fowl | 34 | 0 | 0 | 34 |
| Number of turkeys | 8 | 0 | 8 | 0 |
| Number of dogs | 100 | 27 | 44 | 29 |
| Number of cats | 31 | 10 | 11 | 10 |
| Number of cattle | 23 | 0 | 0 | 74 |
| Number of pigs | 17 | 0 | 20 | 55 |
| Number of goats | 419 | 0 | 48 | 371 |
| **Husbandry characteristics** | n (%) | | | |
| **Where are animals kept?** | | | | |
| **Chickens** | | | | |
| In the house | 70 (60.3%) | 15 (83.3%) | 25 (64.1%) | 30 (50.8%) |
| Shelter /Boma within household compound | 33 (28.4%) | 2 (11.1%) | 12 (30.8%) | 19 (32.2%) |
| Shelter /Boma outside the household compound | 9 (7.8%) | 1 (5.6%) | 1 (2.6%) | 7 (11.9%) |
| Other | 4 (3.4%) | 0 (0.0%) | 1 (2.6%) | 3 (5.1%) |
| **Dogs** | | | | |
| Free roaming | 34 (79.1%) | 9 (64.3%) | 15 (78.9%) | 10 (100.0%) |
| Shelter /Boma outside the household compound | 1 (2.3%) | 1 (7.1%) | 0 (0.0%) | 0 (0.0%) |
| Shelter /Boma within household compound | 8 (18.6%) | 4 (28.6%) | 4 (21.1%) | 0 (0.0%) |
| **Cattle** | | | | |
| Shelter /Boma within household compound | 11 (47.8%) | NA | NA | 11 (47.8%) |
| Shelter /Boma outside the household compound | 11 (47.8%) | NA | NA | 11 (47.8%) |
| Other | 1 (4.4%) | NA | NA | 1 (4.2%) |
| **Goats** | | | | |
| In the house | 8 (16.3%) | NA | 5 (41.7%) | 3 (8.1%) |
| Shelter /Boma within household compound | 28 (57.1%) | NA | 6 (50.0%) | 22 (59.5%) |
| Shelter /Boma outside the household compound | 12 (24.5%) | NA | 1 (8.3%) | 11 (29.7%) |
| Free roaming | 1 (2.0%) | NA | 0 (0.0%) | 1 (2.7%) |
| **Pigs** | | | | |
| Shelter /Boma within household compound | 10 (58.8%) | NA | 5 (100.0%) | 5 (41.7%) |
| Shelter /Boma outside the household compound | 6 (35.3%) | NA | 0 (0.0%) | 6 (50.0%) |
| Free roaming | 1 (5.9%) | NA | 0 (0.0%) | 1 (8.3%) |

| Livestock production system | | | | |
|---|---|---|---|---|
| **Beef cattle** | | | | |
| Zero Grazing | 2 (%) | NA | NA | 2 (10.0%) |
| Communal Grazing | 15 (%) | NA | NA | 15 (75.0%) |
| Pastoral | 3 (%) | NA | NA | 3 (15.0%) |
| **Dairy cattle** | | | | |
| Pastoral | 4 (%) | NA | NA | 4 (100.0%) |
| **Small ruminants** | | | | |
| Zero Grazing | 8 (%) | NA | 6 (50.0%) | 2 (5.4%) |
| Communal Grazing | 26 (%) | NA | 6 (50.0%) | 20 (54.1%) |
| Pastoral | 15 (%) | NA | 0 (0.0%) | 15 (40.5%) |

**S6 Table.** Healthcare choices for household animals

| Animal | Site | Response to sickness in household animals: Consult a governmental veterinarian | Consult a private veterinarian | Use medication from a veterinarian drug store | Use left-over or vet applied drugs | Get medications from friends or family | Use traditional medication | Kill animal (+/- eaten) | Nothing |
|---|---|---|---|---|---|---|---|---|---|
| **Cattle** | Urban | NA | NA | NA | NA | NA | NA | NA | NA |
| | Peri-urban | NA | NA | NA | NA | NA | NA | NA | NA |
| | Rural | n=6 | n=1 | n=1 | n=1 | n=1 | n=0 | n=0 | n=9 |
| **Goats** | Urban | NA | NA | NA | NA | NA | NA | NA | NA |
| | Peri-urban | n=2 | n=1 | n=0 | n=1 | n=0 | n=4 | n=0 | n=3 |
| | Rural | n=9 | n=2 | n=4 | n=2 | n=0 | n=1 | n=0 | n=9 |
| **Pigs** | Urban | NA | NA | NA | NA | NA | NA | NA | NA |
| | Peri-urban | n=1 | n=1 | n=1 | n=0 | n=0 | n=0 | n=0 | n=1 |
| | Rural | n=2 | n=0 | n=1 | n=0 | n=0 | n=1 | n=0 | n=6 |
| **Poultry** | Urban | n=0 | n=1 | n=3 | n=0 | n=0 | n=3 | n=1 | n=6 |
| | Peri-urban | n=7 | n=1 | n=4 | n=2 | n=0 | n=8 | n=1 | n=6 |
| | Rural | n=7 | n=2 | n=7 | n=2 | n=0 | n=5 | n=5 | n=17 |
| **All animals** | **All regions** | n=34 (21.4%) | n=9 (5.6%) | n=21 (13.2%) | n=8 (5.0%) | n=1 (0.6%) | n=22 (13.8%) | n=7 (4.4%) | n=57 (35.8%) |

*^ Yellow = selected by ≥1 households in region. White = not selected. Grey = NA.*

**S7 Table.** Numbers of samples screened for ESBL *E. coli* and ESBL *K. pneumoniae*, stratified by sample type and region.

| Broad sample type | | Sample number n (%): Total | Urban | Peri-urban | Rural |
|---|---|---|---|---|---|
| Human stool | | 2845 (23.8%) | 821 (22.3%) | 982 (24.4%) | 1042 (24.3%) |
| Animal stool | | 973 (8.1%) | 118 (3.2%) | 229 (5.7%) | 626 (14.6%) |
| Environment | | 8157 (68.1%) | 2736 (74.5%) | 2807 (69.9%) | 2614 (60.1%) |
| | Food | 1168 (9.8%) | 333 (9.1%) | 440 (11.0%) | 395 (9.2%) |
| | Drinking water | 1254 (10.5%) | 532 (14.5%) | 449 (11.2%) | 273 (6.4%) |
| | Source water | 527 (4.4%) | 79 (2.1%) | 216 (5.4%) | 232 (5.4%) |
| | Household surfaces | 2458 (20.5%) | 766 (20.8%) | 744 (18.5%) | 948 (22.1%) |
| | Household floor | 745 (6.2%) | 247 (6.7%) | 244 (6.1%) | 254 (5.9%) |
| | Clothing | 742 (6.2%) | 245 (6.7%) | 242 (6.0%) | 255 (5.9%) |
| | Hand-contact samples | 451 (3.8%) | 129 (3.5%) | 69 (1.7%) | 253 (5.9%) |
| | Household drains | 300 (2.5%) | 151 (4.1%) | 149 (3.7%) | n=0 (0.0%) |
| | River water | 512 (4.3%) | 254 (6.9%) | 254 (6.3%) | 4 (0.1%) |
| **TOTAL** | | **11975** | **3675** | **4018** | **4282** |

**S8a Table.** Regional univariate analysis of WASH and individual variables against human ESBL *E. coli* colonisation

| Characteristic | Region | n | OR | 95% CI | p value | Model inclusion |
|---|---|---|---|---|---|---|
| **Season (wet)** | Urban | 813 | 1.11 | 0.84,1.47 | 0.5 | **Yes** |
| | Peri | 971 | **1.92** | **1.47,2.53** | **<0.001** | |
| | Rural | 938 | 1.29 | 0.98,1.69 | 0.067 | |
| **Male sex** | Urban | 813 | 0.82 | 0.61,1.09 | 0.2 | No |
| | Peri | 971 | 0.93 | 0.71,1.22 | 0.6 | |
| | Rural | 938 | 0.97 | 0.66,1.14 | 0.3 | |
| **Age (log)** | Urban | 813 | **1.14** | **1.01,1.29** | **0.030** | **Yes** |
| | Peri | 971 | 1.07 | 0.94,1.21 | 0.3 | |
| | Rural | 938 | 1.07 | 0.95,1.21 | 0.3 | |
| **ABU** *(Last 6 months)* | Urban | 813 | 1.01 | 0.71,1.45 | >0.9 | No |
| | Peri | 971 | 0.89 | 0.57,1.37 | 0.6 | |
| | Rural | 938 | 1.15 | 0.83,1.57 | 0.4 | |
| **HIV reactive** | Urban | 813 | 0.85 | 0.48,1.49 | 0.6 | No |
| | Peri | 971 | 0.86 | 0.49,1.47 | 0.6 | |
| | Rural | 938 | 1.23 | 0.77,1.94 | 0.4 | |
| **Household density (log)** | Urban | 813 | 1.17 | 0.82,1.67 | 0.4 | **Yes** |
| | Peri | 971 | 0.99 | 0.71,1.39 | >0.9 | |
| | Rural | 938 | **0.66** | **0.45,0.97** | **0.034** | |
| **Income** *(>40,000MK/month)* | Urban | 813 | 0.91 | 0.69,1.21 | 0.5 | No |
| | Peri | 971 | 1.04 | 0.8,1.37 | 0.8 | |
| | Rural | 938 | 0.84 | 0.64,1.11 | 0.2 | |
| **Shared Toilet** | Urban | 813 | 1.02 | 0.77,1.35 | 0.9 | **Yes** |
| | Peri | 971 | **1.37** | **1.04, 1.81** | **0.026** | |
| | Rural | 938 | 0.86 | 0.63,1.18 | 0.4 | |
| **Drophole Present** | Urban | 813 | 0.80 | 0.55, 1.15 | 0.2 | **Yes** |
| | Peri | 971 | **0.59** | **0.44,0.79** | **<0.001** | |
| | Rural | 938 | 1.08 | 0.81,1.44 | 0.6 | |
| **Cleaning Materials available** | Urban | 813 | 1.17 | 0.84,1.62 | 0.3 | **Yes** |
| | Peri | 971 | **0.65** | **0.49,0.84** | **0.001** | |
| | Rural | 938 | 0.80 | 0.51,1.23 | 0.3 | |
| **Human Feaces visible** | Urban | 813 | 0.95 | 0.72,1.25 | 0.7 | **Yes** |
| | Peri | 971 | **1.47** | **1.08.2.01** | **0.015** | |
| | Rural | 938 | 0.82 | 0.63,1.08 | 0.2 | |
| **Human defecation practiced** | Urban | 813 | 1.47 | 0.71,3.04 | 0.3 | **Yes** |
| | Peri | 971 | 1.02 | 0.68,1.52 | >0.9 | |
| | Rural | 938 | **0.59** | **0.37,0.91** | **0.021** | |
| **HWF present** | Urban | 813 | 1.15 | 0.87,1.52 | 0.3 | No |
| | Peri | 971 | 0.74 | 0.53,1.06 | 0.1 | |
| | Rural | 938 | 1.07 | 0.82,1.40 | 0.6 | |
| **Soap present** | Urban | 813 | **1.89** | **1.17,3.08** | **0.010** | **Yes** |
| | Peri | 971 | **0.74** | **0.56,0.98** | **0.034** | |
| | Rural | 938 | **0.47** | **0.21,0.94** | **0.044** | |
| **Stored water covered** | Urban | 813 | **2.16** | **1.38,3.47** | **<0.001** | **Yes** |

| | | | | | | |
|---|---|---|---|---|---|---|
| | Peri | 971 | 1.30 | 0.97,1.75 | 0.082 | |
| | Rural | 938 | **0.62** | **0.46,0.81** | **<0.001** | |
| **Stored water covered and tap** | Urban | 813 | 0.83 | 0.62,1.10 | 0.2 | No |
| | Peri | 971 | 0.76 | 0.46,1.21 | 0.3 | |
| | Rural | 938 | 0.76 | 0.57,1.01 | 0.056 | |
| **Utensil water** | Urban | 813 | 0.95 | 0.71,1.27 | 0.7 | No |
| | Peri | 971 | 1.43 | 0.93,2.18 | 0.10 | |
| | Rural | 938 | 1.22 | 0.84,1.74 | 0.3 | |
| **Piped water (i.e. kiosk)** | Urban | 813 | 0.97 | 0.73,1.29 | 0.9 | **Yes** |
| | Peri | 971 | **0.64** | **0.47,0.88** | **0.006** | |
| | Rural | 938 | **0.68** | **0.47,0.97** | **0.036** | |
| **Tap water (i.e. household tap)** | Urban | 813 | 0.87 | 0.66,1.15 | 0.3 | **Yes** |
| | Peri | 971 | 0.98 | 0.64,1.50 | >0.9 | |
| | Rural | 938 | **2.38** | **1.19,4.86** | **0.015** | |
| **Tube well water** | Urban | 813 | **2.53** | **1.45,4.54** | **0.001** | **Yes** |
| | Peri | 971 | **1.37** | **1.04,1.82** | **0.027** | |
| | Rural | 938 | 1.31 | 0.93,1.86 | 0.12 | |
| **Animal owned by household** | Urban | 813 | **1.55** | **1.17,2.06** | **0.002** | **Yes** |
| | Peri | 971 | 0.99 | 0.75,1.30 | >0.9 | |
| | Rural | 938 | **0.54** | **0.37,0.78** | **0.001** | |
| **Cattle or ruminant owned** | Urban | 813 | NA | NA | NA | No |
| | Peri | 971 | 2.33 | 1.64,1.11 | 0.2 | |
| | Rural | 938 | 0.89 | 0.68,1.17 | 0.4 | |
| **Poultry owned** | Urban | 813 | **1.61** | **1.15,2.26** | **0.005** | **Yes** |
| | Peri | 971 | 0.85 | 0.64,1.11 | 0.2 | |
| | Rural | 938 | 0.98 | 0.72,1.32 | 0.9 | |
| **Pet owned** | Urban | 813 | 1.24 | 0.90,1.69 | 0.2 | **Yes** |
| | Peri | 971 | 0.85 | 0.62,1.17 | 0.3 | |
| | Rural | 938 | **0.66** | **0.48,0.91** | **0.012** | |
| **Pig owned** | Urban | 813 | NA | NA | NA | **Yes** |
| | Peri | 971 | **0.30** | **0.09,0.78** | **0.026** | |
| | Rural | 938 | 0.93 | 0.66,1.31 | 0.7 | |
| **Animal kept inside house** | Urban | 813 | **1.50** | **1.05,2.15** | **0.024** | **Yes** |
| | Peri | 971 | 1.26 | 0.94,1.67 | 0.12 | |
| | Rural | 938 | **1.16** | **1.18,2.19** | **0.003** | |
| **Animal interacting with food** | Urban | 813 | **1.69** | **1.21,2.36** | **0.002** | **Yes** |
| | Peri | 971 | **1.38** | **1.05,1.81** | **0.020** | |
| | Rural | 938 | **1.41** | **1.07,1.85** | **0.014** | |
| **Animal faeces seen** | Urban | 813 | 1.02 | 0.77,1.34 | >0.9 | No |
| | Peri | 971 | 1.08 | 0.77,1.52 | 0.7 | |
| | Rural | 938 | NA | NA | NA | |
| **River water exposure** | Urban | 813 | 0.89 | 0.65,1.21 | 0.5 | **Yes** |
| | Peri | 971 | 1.33 | 1.00,1.78 | 0.054 | |
| | Rural | 938 | **1.61** | **1.18,2.19** | **0.003** | |
| **Drain water exposure** | Urban | 813 | 0.76 | 0.44,1.29 | 0.3 | **Yes** |
| | Peri | 971 | 1.30 | 0.86,1.94 | 0.2 | |
| | Rural | 938 | **0.49** | **0.27,0.84** | **0.013** | |
| **Street food use** | Urban | 813 | 0.51 | 0.33,1.09 | 0.2 | **Yes** |

| | | | | | | |
|---|---|---|---|---|---|---|
| | Peri | 971 | 1.56 | 1.01,2.49 | 0.053 | |
| | Rural | 938 | **1.65** | **1.19,2.29** | **0.003** | |
| **Shared plates** | Urban | 813 | 0.79 | 0.58,1.09 | 0.2 | **Yes** |
| | Peri | 971 | 1.12 | 0.85,1.47 | 0.4 | |
| | Rural | 938 | **0.71** | **0.54,0.94** | **0.016** | |
| **Market produce used** | Urban | 813 | 0.65 | 0.40,1.05 | 0.08 | No |
| | Peri | 971 | 0.69 | 0.45,1.05 | 0.078 | |
| | Rural | 938 | 0.95 | 0.72,1.26 | 0.7 | |

**S8b Table.** Regional univariate analysis of WASH and individual variables against human ESBL *K. pnuemoniae* colonisation

| Characteristic | Region | n | OR | 95% CI | p value | Model inclusion |
|---|---|---|---|---|---|---|
| **Season (wet)** | Urban | 813 | 1.30 | 0.85,2.00 | 0.2 | **Yes** |
| | Peri | 971 | **1.94** | **1.27,3.02** | **0.003** | |
| | Rural | 938 | **2.19** | **1.47,3.31** | **<0.001** | |
| **Male sex** | Urban | 813 | 0.69 | 0.43,1.09 | 0.12 | No |
| | Peri | 971 | 1.12 | 0.74,1.68 | 0.6 | |
| | Rural | 938 | 0.84 | 0.56,1.23 | 0.4 | |
| **Age (log)** | Urban | 813 | 0.97 | 0.82,1.17 | 0.8 | No |
| | Peri | 971 | 0.98 | 0.82,2.60 | 0.2 | |
| | Rural | 938 | 1.02 | 0.86,1.21 | 0.8 | |
| **ABU** *(Last 6 months)* | Urban | 813 | 1.28 | 0.74,2.11 | 0.4 | **Yes** |
| | Peri | 971 | 0.75 | 0.34,1.46 | 0.4 | |
| | Rural | 938 | **1.54** | **0.99,2.35** | **0.048** | |
| **HIV reactive** | Urban | 813 | 1.12 | 0.45,2.41 | 0.8 | **Yes** |
| | Peri | 971 | **2.29** | **1.15,4.24** | **0.012** | |
| | Rural | 938 | 0.52 | 0.20,1.13 | 0.14 | |
| **Household density (log)** | Urban | 813 | **2.29** | **1.32,3.99** | **0.003** | **Yes** |
| | Peri | 971 | 0.95 | 0.57,1.60 | 0.8 | |
| | Rural | 938 | **2.12** | **1.24,3.60** | **0.006** | |
| **Income** *(>40,000MK/month)* | Urban | 813 | 1.16 | 0.76,1.78 | 0.5 | No |
| | Peri | 971 | 1.10 | 0.73,1.67 | 0.7 | |
| | Rural | 938 | 1.04 | 0.71,1.54 | 0.8 | |
| **Shared Toilet** | Urban | 813 | 1.39 | 0.91,2.16 | 0.13 | No |
| | Peri | 971 | 0.87 | 0.55, 1.34 | 0.5 | |
| | Rural | 938 | 1.32 | 0.85,2.00 | 0.2 | |
| **Drophole Present** | Urban | 813 | 1.07 | 0.60,1.81 | 0.8 | No |
| | Peri | 971 | 0.96 | 0.61, 1.47 | 0.9 | |
| | Rural | 938 | 1.33 | 0.89, 1.98 | 0.2 | |
| **Cleaning Materials available** | Urban | 813 | 1.07 | 0.64,1.73 | 0.8 | No |
| | Peri | 971 | 1.06 | 0.70,1.59 | 0.8 | |
| | Rural | 938 | 0.78 | 0.39,1.45 | 0.5 | |
| **Human Feaces visible** | Urban | 813 | **1.58** | **1.03,2.44** | **0.039** | **Yes** |
| | Peri | 971 | 1.44 | 0.90,2.25 | 0.12 | |
| | Rural | 938 | 1.01 | 0.68,1.49 | >0.9 | |

| | | | | | | |
|---|---|---|---|---|---|---|
| **Human defecation practiced** | Urban | 813 | 0.79 | 0.19,2.30 | 0.7 | No |
| | Peri | 971 | 0.75 | 0.36,1.41 | 0.4 | |
| | Rural | 938 | 0.64 | 0.31,1.21 | 0.2 | |
| **HWF present** | Urban | 813 | 1.55 | 1.00,2.42 | 0.053 | No |
| | Peri | 971 | 1.02 | 0.60,1.86 | >0.9 | |
| | Rural | 938 | 1.29 | 0.88,1.93 | 0.2 | |
| **Soap present** | Urban | 813 | 1.19 | 0.56,2.30 | 0.6 | No |
| | Peri | 971 | 0.72 | 0.46,1.10 | 0.13 | |
| | Rural | 938 | 0.31 | 0.05,1.04 | 0.11 | |
| **Stored water covered** | Urban | 813 | 0.83 | 0.46,1.58 | 0.5 | No |
| | Peri | 971 | 0.73 | 0.48,1.12 | 0.14 | |
| | Rural | 938 | 1.08 | 0.73,1.60 | 0.7 | |
| **Stored water covered and tap** | Urban | 813 | 0.84 | 0.54,1.31 | 0.5 | No |
| | Peri | 971 | 0.68 | 0.28,1.42 | 0.3 | |
| | Rural | 938 | 1.06 | 0.71,1.58 | 0.8 | |
| **Utensil water** | Urban | 813 | 0.80 | 0.50,1.26 | 0.3 | No |
| | Peri | 971 | 0.62 | 0.25,1.28 | 0.2 | |
| | Rural | 938 | 0.88 | 0.49,1.48 | 0.6 | |
| **Piped water (i.e. kiosk)** | Urban | 813 | 1.53 | 1.00,2.34 | 0.052 | No |
| | Peri | 971 | 0.85 | 0.52,1.36 | 0.5 | |
| | Rural | 938 | 1.36 | 0.84,2.15 | 0.2 | |
| **Tap water (i.e. household tap)** | Urban | 813 | 0.66 | 0.43,1.02 | 0.061 | No |
| | Peri | 971 | 1.31 | 0.69,2.33 | 0.4 | |
| | Rural | 938 | 0.21 | 0.01, 1.00 | 0.13 | |
| **Tube well water** | Urban | 813 | 0.91 | 0.34,2.03 | 0.8 | No |
| | Peri | 971 | 1.08 | 0.71,1.67 | 0.7 | |
| | Rural | 938 | 0.80 | 0.51, 1.28 | 0.3 | |
| **Animal owned by household** | Urban | 813 | 1.39 | 0.91,2.13 | 0.13 | No |
| | Peri | 971 | 0.81 | 0.53,1.22 | 0.3 | |
| | Rural | 938 | 0.70 | 0.43, 1.20 | 0.2 | |
| **Cattle or ruminant owned** | Urban | 813 | NA | NA | NA | No |
| | Peri | 971 | 1.06 | 0.59,1.80 | 0.8 | |
| | Rural | 938 | 1.22 | 0.83,1.80 | 0.3 | |
| **Poultry owned** | Urban | 813 | **1.46** | **1.12,2.87** | **0.013** | **Yes** |
| | Peri | 971 | 1.12 | 0.74,1.69 | 0.6 | |
| | Rural | 938 | 1.37 | 0.88,2.21 | 0.2 | |
| **Pet owned** | Urban | 813 | 1.33 | 0.55,1.47 | 0.7 | **Yes** |
| | Peri | 971 | 1.01 | 0.61,1.62 | >0.9 | |
| | Rural | 938 | **1.59** | **1.05,2.39** | **0.027** | |
| **Pig owned** | Urban | 813 | NA | NA | NA | No |
| | Peri | 971 | NA | NA | NA | |
| | Rural | 938 | 1.56 | 0.98,2.42 | 0.055 | |
| **Animal kept inside house** | Urban | 813 | 1.46 | 0.87,2.40 | 0.14 | **Yes** |
| | Peri | 971 | 0.52 | 0.30,0.86 | **0.014** | |
| | Rural | 938 | **1.48** | **1.00,2.19** | **0.048** | |
| **Animal interacting with food** | Urban | 813 | 1.33 | 0.80,2.15 | 0.3 | No |
| | Peri | 971 | 0.92 | 0.60,1.40 | 0.7 | |
| | Rural | 938 | 0.69 | 0.46,1.01 | 0.058 | |

| | | | | | | |
|---|---|---|---|---|---|---|
| **Animal faeces seen** | Urban | 813 | **0.63** | **0.40,0.96** | **0.036** | **Yes** |
| | Peri | 971 | 1.42 | 0.82,2.60 | 0.2 | |
| | Rural | 938 | NA | NA | NA | |
| **River water exposure** | Urban | 813 | 1.48 | 0.94,2.30 | 0.088 | No |
| | Peri | 971 | 1.28 | 0.82,2.04 | 0.3 | |
| | Rural | 938 | 0.81 | 0.49,1.28 | >0.9 | |
| **Drain water exposure** | Urban | 813 | 1.46 | 0.68,2.85 | 0.3 | **Yes** |
| | Peri | 971 | 0.91 | 0.45,1.68 | 0.8 | |
| | Rural | 938 | **2.82** | **1.59,4.83** | **<0.001** | |
| **Street food use** | Urban | 813 | 0.52 | 0.30,0.92 | **0.036** | **Yes** |
| | Peri | 971 | 0.86 | 0.48,1.67 | 0.6 | |
| | Rural | 938 | 0.90 | 0.59,1.41 | 0.6 | |
| **Shared plates** | Urban | 813 | 0.71 | 0.42,1.16 | 0.2 | **Yes** |
| | Peri | 971 | 0.84 | 0.54,1.27 | 0.4 | |
| | Rural | 938 | **0.64** | **0.43,0.95** | **0.025** | |
| **Market produce used** | Urban | 813 | 1.13 | 0.56,2.63 | 0.7 | No |
| | Peri | 971 | 0.80 | 0.44,1.55 | 0.5 | |
| | Rural | 938 | 1.13 | 0.75,1.71 | 0.6 | |

**S9a Table.** Table of parameter testing for regional adjustment of variables included in the ESBL *E. coli* mixed effects model

| **Characteristic** | **Likelihood ratio test** | **Adjust for Region*** |
|---|---|---|
| **Season** | $\chi 2$ (2)= 8.33, $p$ = 0.0155 | **Yes** |
| **Male** | NA | |
| **Age** | $\chi 2$ (2)= 0.80, $p$ =0.67 | No |
| **ABU** | NA | |
| **HIV reactive** | NA | |
| **Household density** | $\chi 2$ (2)= 4.82, $p$ =0.090 | No |
| **Income >40,000MK/month** | NA | |
| **Shared Toilet** | $\chi 2$ (2)= 4.93, $p$ = 0.085 | No |
| **Drophole Present** | $\chi 2$ (2)= 8.51, $p$ = 0.014 | **Yes** |
| **Cleaning Materials available** | $\chi 2$ (2)= 7.66, $p$ = 0.022 | **Yes** |
| **Human Feaces visible** | $\chi 2$ (2)= 7.88, $p$ = 0.019 | **Yes** |
| **Human defecation practiced** | $\chi 2$ (2)= 5.64, $p$ = 0.059 | No |
| **HWF present** | NA | |
| **Soap present** | $\chi 2$ (2)= 14.32, $p$ = <0.001 | **Yes** |
| **Stored water covered** | $\chi 2$ (2)= 26.52, $p$ = <0.001 | **Yes** |
| **Stored water covered and tap** | NA | |
| **Utensil water** | NA | |
| **Piped water (i.e. kiosk)** | $\chi 2$ (2)= 4.39, $p$ =0.111 | No |
| **Tap water (i.e. household tap)** | $\chi 2$ (2)= 6.92, $p$ =0.031 | **Yes** |
| **Tube well water** | $\chi 2$ (2)= 4.28, $p$ =0.117 | No |

| | | |
|---|---|---|
| **Animal owned by household** | $\chi 2$ (2)= 19.61, $p$ = <0.001 | **Yes** |
| **Cattle or ruminant owned** | NA | |
| **Poultry owned** | $\chi 2$ (2)= 8.91, $p$ =0.011 | **Yes** |
| **Pet owned** | $\chi 2$ (2)= 7.63, $p$ =0.022 | **Yes** |
| **Pig owned** | NA | |
| **Animal kept inside house** | $\chi 2$ (2)= 1.34, $p$ = 0.510 | No |
| **Animal interacting with food** | $\chi 2$ (2)= 0.94, $p$ = 0.624 | No |
| **Animal faeces seen** | NA | |
| **River water exposure** | $\chi 2$ (2)= 7.36, $p$ = 0.025 | **Yes** |
| **Drain water exposure** | $\chi 2$ (2)= 8.23, $p$ = 0.016 | **Yes** |
| **Street food use** | $\chi 2$ (2)= 20.84, $p$ = <0.001 | **Yes** |
| **Shared plates** | $\chi 2$ (2)= 5.72, $p$ = 0.057 | No |
| **Market produce used** | NA | |

**An alpha level 0.05 has been used as a cut off for the decision to adjust for regional effects in the final mixed effect model.*

**S9b Table.** Table of parameter testing for regional adjustment of variables included in the ESBL *K. pnuemoniae* mixed effects model

| Variable | Likelihood ratio test | Adjust for Region* |
|---|---|---|
| **Season** | $\chi 2$ (2)= 3.25, $p$ =0.197 | No |
| **Male** | NA | |
| **Age** | NA | No |
| **ABU** | $\chi 2$ (2)= 3.08, $p$ =0.215 | No |
| **HIV reactive** | $\chi 2$ (2)= 7.87, $p$ =0.020 | **Yes** |
| **Household density** | $\chi 2$ (2)= 6.53 , $p$ = 0.038 | **Yes** |
| **Income >40,000MK/month** | NA | |
| **Shared Toilet** | NA | |
| **Drophole Present** | NA | |
| **Cleaning Materials available** | NA | |
| **Human Feaces visible** | $\chi 2$ (2)= 2.56, $p$ =0.278 | No |
| **Human defecation practiced** | NA | |
| **HWF present** | NA | |
| **Soap present** | NA | |
| **Stored water covered** | NA | |
| **Stored water covered and tap** | NA | |
| **Utensil water** | NA | |
| **Piped water (i.e. kiosk)** | NA | |
| **Tap water (i.e. household tap)** | NA | |
| **Tube well water** | NA | |
| **Animal owned by household** | NA | |
| **Cattle or ruminant owned** | NA | |

| | | |
|---|---|---|
| **Poultry owned** | $\chi 2$ (2)= 2.24, $p$ =0.327 | No |
| **Pet owned** | $\chi 2$ (2)= 3.46, $p$ =0.177 | No |
| **Pig owned** | NA | |
| **Animal kept inside house** | $\chi 2$ (2)= 12.39, $p$ =0.002 | **Yes** |
| **Animal interacting with food** | NA | |
| **Animal faeces seen** | $\chi 2$ (2)= 5.56, $p$ =0.062 | No |
| **River water exposure** | NA | |
| **Drain water exposure** | $\chi 2$ (2)= 7.02, $p$ =0.030 | **Yes** |
| **Street food use** | $\chi 2$ (2)= 2.54, $p$ =0.281 | No |
| **Shared plates** | $\chi 2$ (2)= 0.81, $p$ =0.665 | No |
| **Market produce used** | NA | |

**An alpha level 0.05 has been used as a cut off for the decision to adjust for regional effects in the final mixed effect model.*

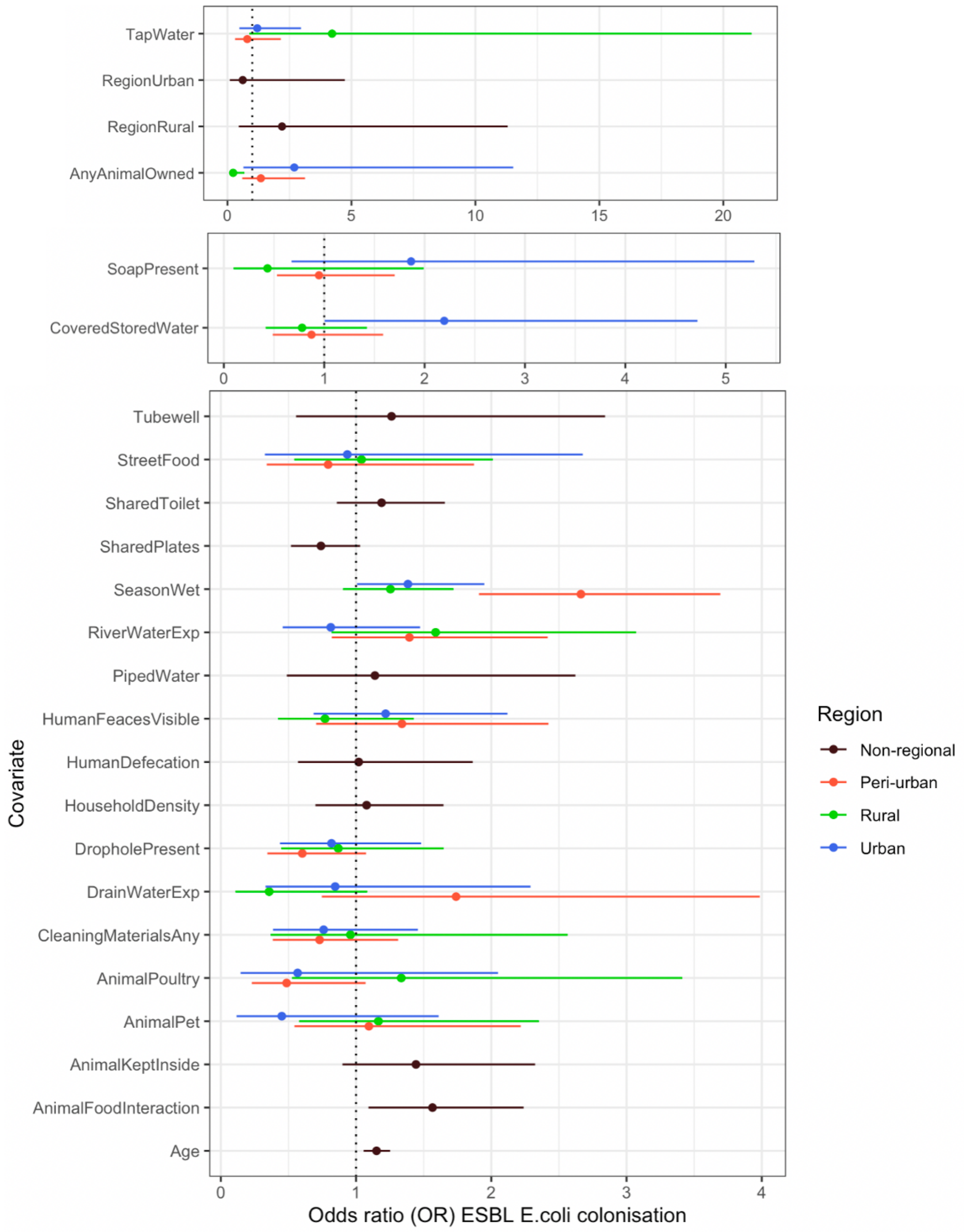


**S1a Fig.** Parameter estimates for the fixed-effects used in a multivariable model of ESBL *E. coli* colonisation, expressed as odds ratios with 95% CrI. Covariates either included an interaction term by region (and so their effect varies by region - red=peri-urban, green=rural or blue=urban) or had the same effect across region (black). **Covariates that were significantly associated (p<0.05) with colonisation via univariable analysis in any region were evaluated for a different effect across regions by comparing models with and without a covariate*region interaction term using likelihood ratio*

*testing for both ESBL E. coli and ESBL K. pneumoniae. An interaction term with region was included for those covariates for which p < 0.05 on likelihood ratio testing (**S7a Table**).*

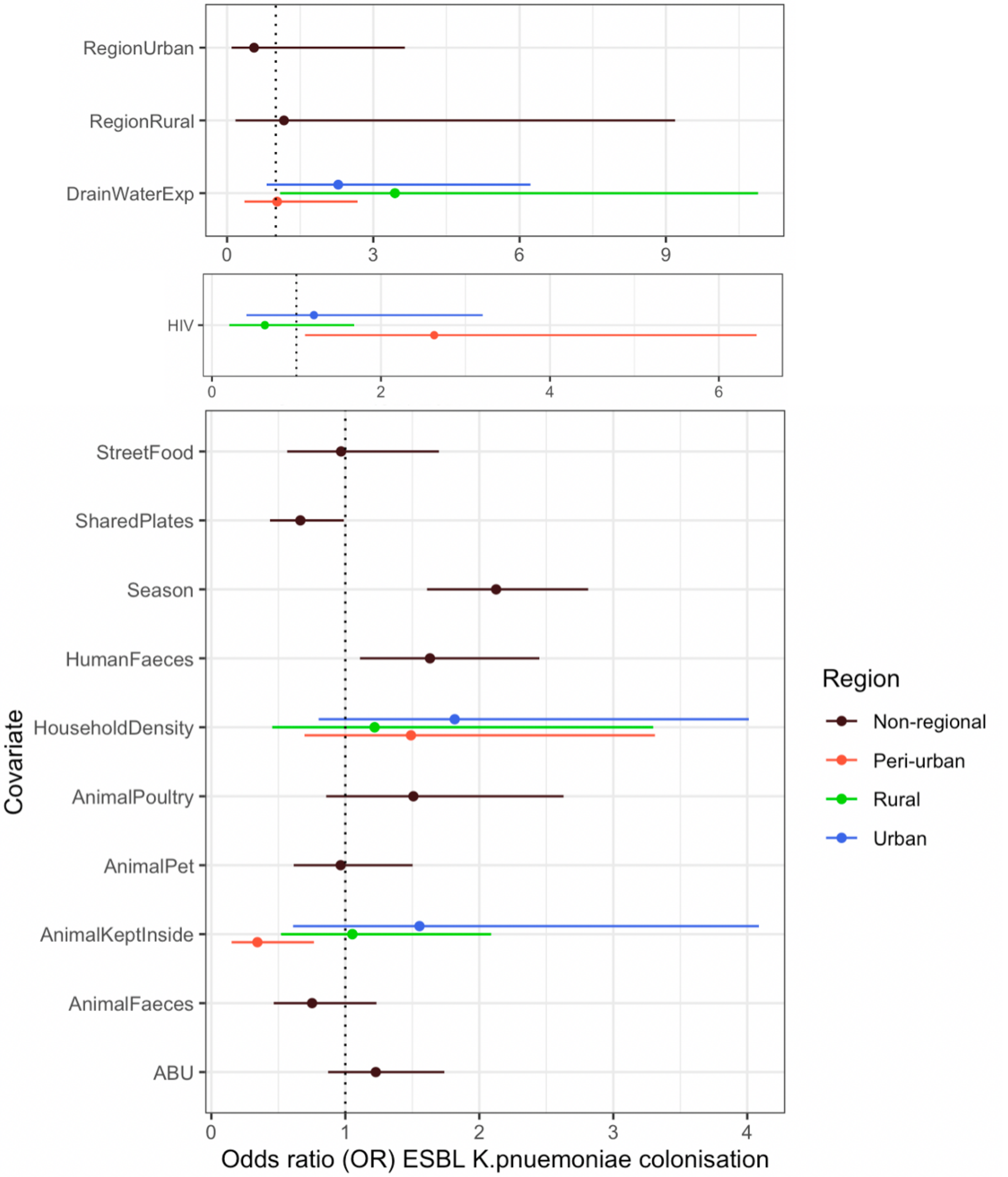


**S1b Fig.** Parameter estimates for the fixed-effects used in a multivariable model of ESBL *K. pneumoniae* colonisation, expressed as odds ratios with 95% CrI. Covariates either included an interaction term by region (and so their effect varies by region - red=peri-urban, green=rural or blue=urban) or had the same effect across regions (black). *Covariates that were significantly associated ($p<0.05$) with colonisation via univariable analysis in any region were evaluated for a different effect across regions by comparing models with and without a covariate*region interaction term using likelihood ratio testing for both ESBL E. coli and ESBL K. pneumoniae. An interaction term with region was included for those covariates for which $p < 0.05$ on likelihood ratio testing (**S7b Table**).*

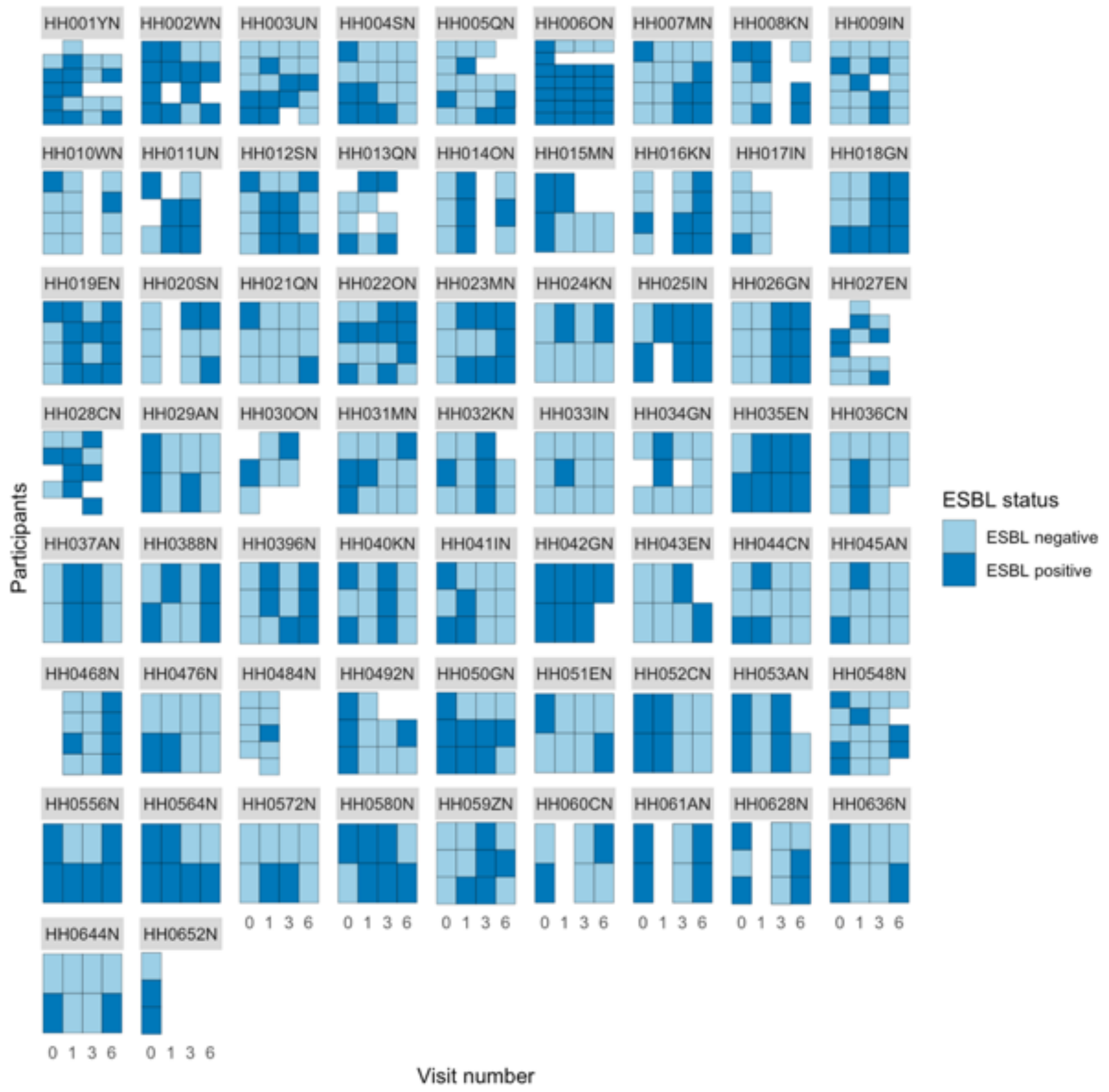


**S2a Fig.** Facet Plot showing flux of human ESBL (*E. coli* or *K. pneumoniae*) colonisation amongst **urban** household members over time, grouped by the 65 households recruited. Each row represents a participant, each column represents a visit, and each small square is a sample coloured by EBSL status (positive or negative). Where no sample was returned for an individual at a visit the square remains blank.

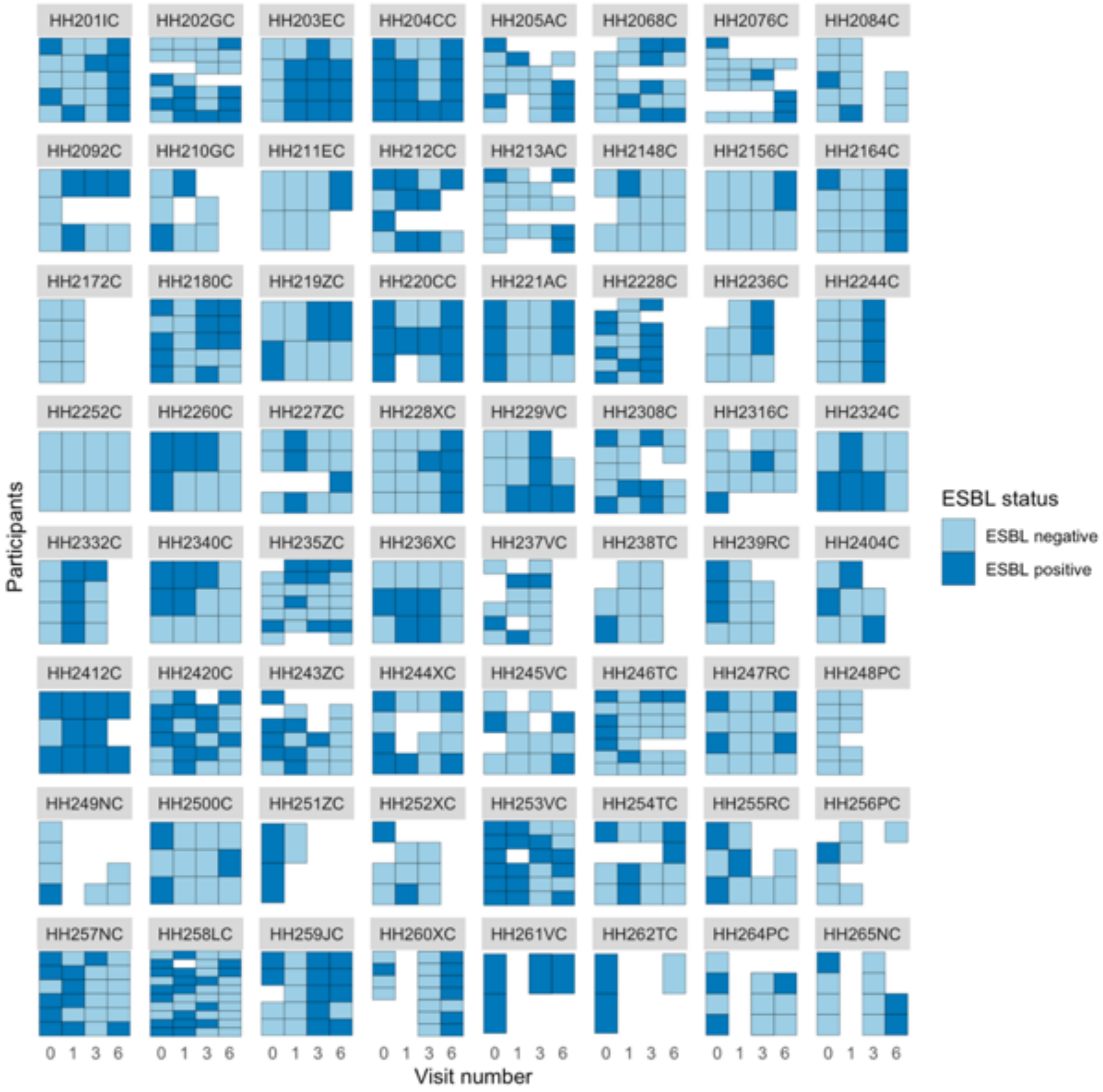


**S2b Fig.** Facet Plot showing flux of human ESBL colonisation (*E. coli* or *K. pneumoniae*) amongst **peri-urban** household members over time, grouped by the 65 households recruited. Each row represents a participant, each column represents a visit, and each small square is a sample coloured by EBSL status (positive or negative). Where no sample was returned for an individual at a visit the square remains blank.

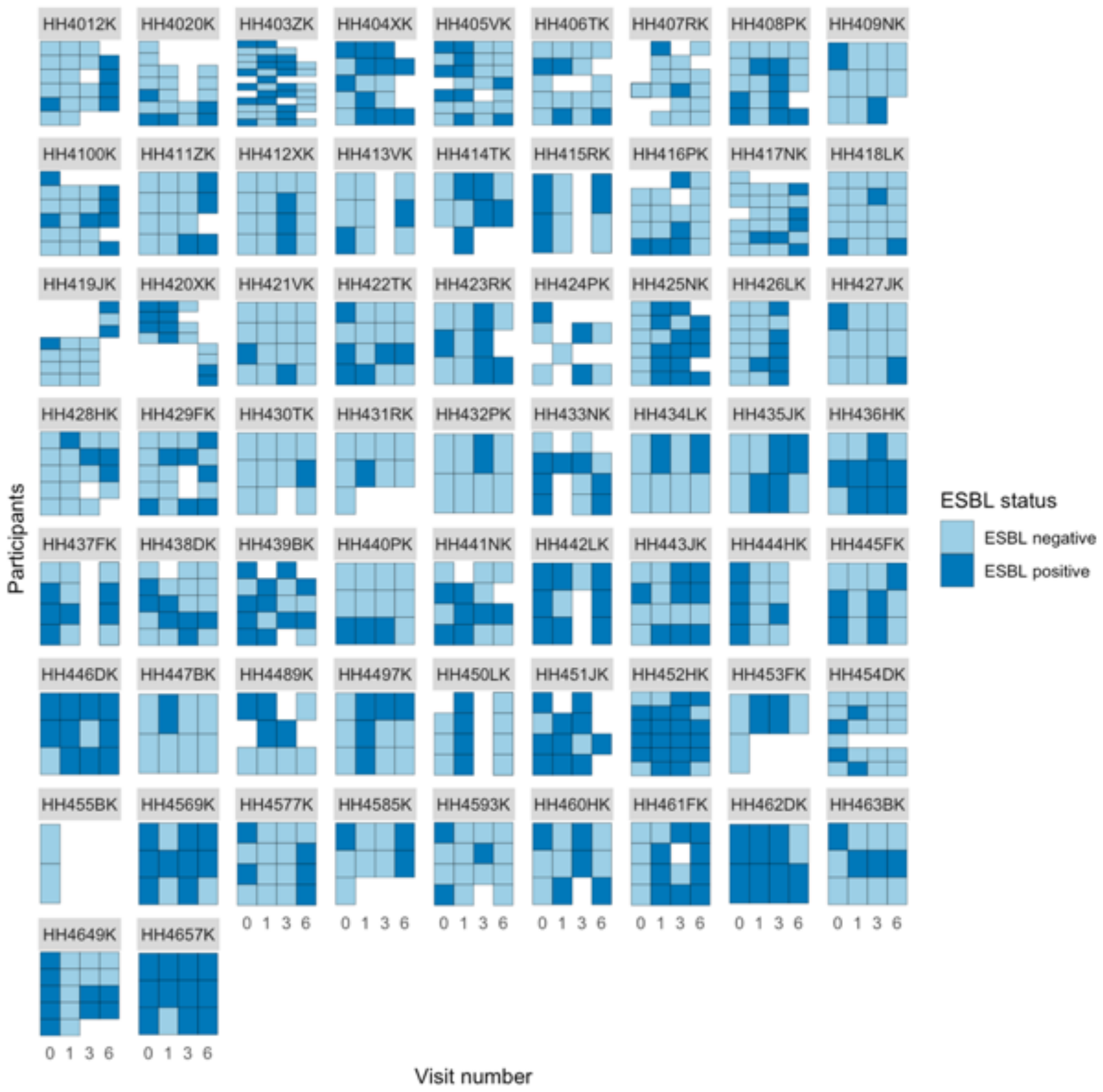


**S2c Fig.** Facet Plot showing flux of human ESBL (*E. coli* or *K. pneumoniae*) colonisation amongst **rural** household members over time, grouped by the 65 households recruited. Each row represents a participant, each column represents a visit, and each small square is a sample coloured by EBSL status (positive or negative). Where no sample was returned for an individual at a visit the square remains blank.

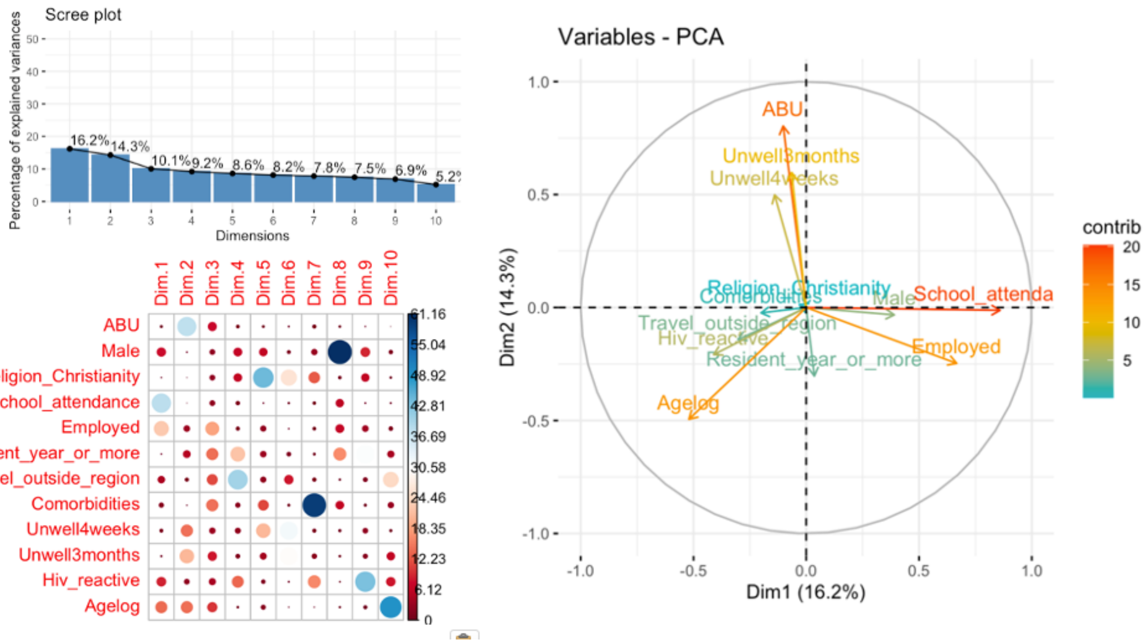


**S3a Fig.** PCA analysis of individual variables, including a scree plot of the eigenvalues (top left), weighting of the variables by PCA (bottom left), and factor map (right).

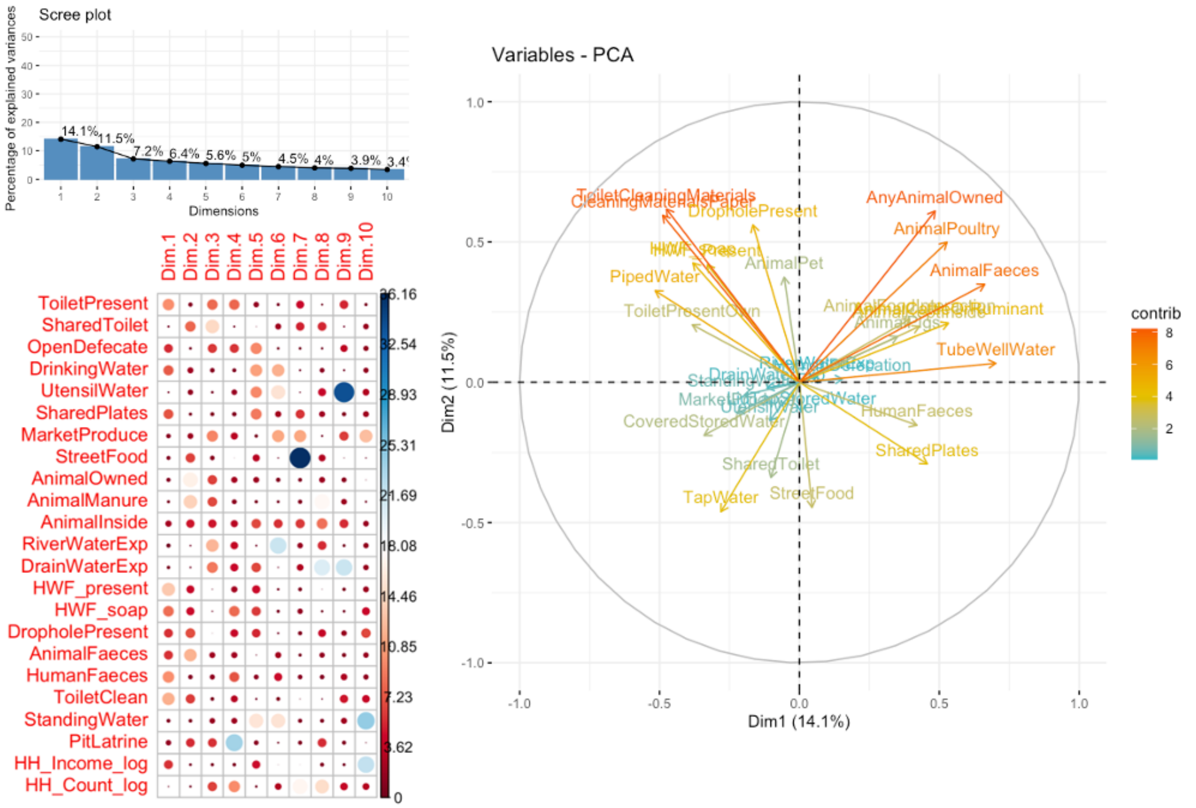


**S3b Fig.** PCA analysis of household variables, including a scree plot of the eigenvalues (top left), weighting of the variables by PCA (bottom left), and factor map (right).

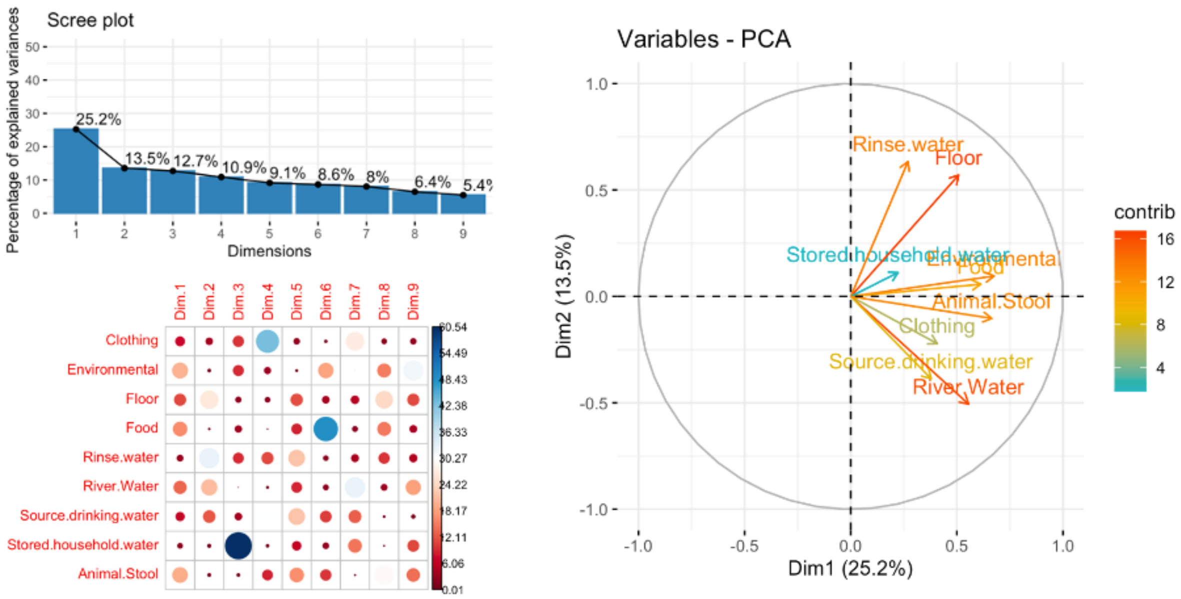


**S3c Fig.** PCA analysis of environmental contamination variables, including a scree plot of the eigenvalues (top left), weighting of the variables by PCA (bottom left), and factor map (right).

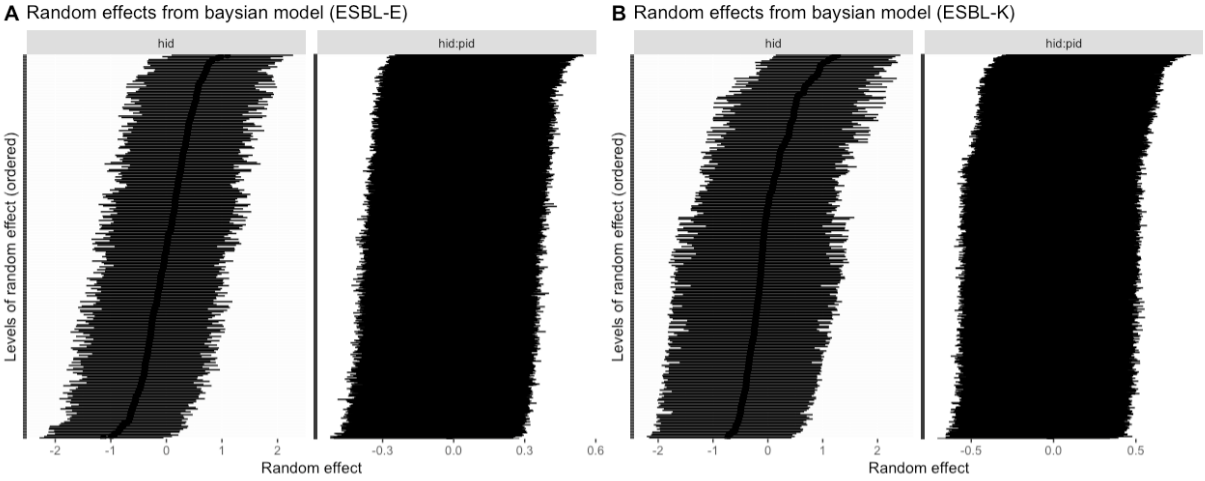


**S4 Fig.** Random effects from Bayesian multivariate models of (a) ESBL *E. coli* [ESBL-E], and (b) ESBL *K. pnuemoniae* [ESBL-K], inclusive of within household (hid) and within participant (hid:pid) effects.
